# SPECIFICATIONS OF THE ACMG/AMP VARIANT CURATION GUIDELINES FOR MYOCILIN: RECOMMENDATIONS FROM THE CLINGEN GLAUCOMA EXPERT PANEL

**DOI:** 10.1101/2022.02.16.22271025

**Authors:** Kathryn P Burdon, Patricia Graham, Johanna Hadler, John D Hulleman, Francesca Pasutto, Erin A Boese, Jamie E Craig, John H Fingert, Alex W Hewitt, Owen M Siggs, Kristina Whisenhunt, Terri L Young, David A Mackey, Andrew Dubowsky, Emmanuelle Souzeau

## Abstract

The standardization of variant curation criteria is essential for accurate interpretation of genetic results and clinical care of patients. The variant curation guidelines developed by the American College of Medical Genetics and Genomics (ACMG) and the Association for Molecular Pathology (AMP) in 2015 are widely used but are not gene specific. To address this issue, the Clinical Genome Resource (ClinGen) Variant Curation Expert Panels (VCEP) have been tasked with developing gene-specific variant curation guidelines. The Glaucoma VCEP was created to develop rule specifications for genes associated with primary glaucoma, including *myocilin* (*MYOC*), the most common cause of Mendelian glaucoma. Of the 28 ACMG/AMP criteria, the Glaucoma VCEP adapted 15 rules to *MYOC*, and deemed 13 rules not applicable. Key specifications included determining minor allele frequency thresholds, developing an approach to counting probands and segregations, and reviewed functional assays. The rules were piloted on 81 variants and led to a change in classification in 38% of those that were classified in ClinVar with functional evidence influencing the classification of 18 variants. The standardized variant curation guidelines for *MYOC* provide a framework for the consistent application of the rules between laboratories, to improve *MYOC* genetic testing in the management of glaucoma.

## INTRODUCTION

Accurately interpreting sequence variants is a key component in genetic diagnosis. However, the abundance of evidence available and the difficulty in objectively assessing and weighing the different types of evidence render the task complex for clinical laboratories.

Harmonized variant curation criteria are essential for accurately interpreting genetic results and the clinical care of patients. In 2015, the American College of Medical Genetics and Genomics (ACMG) and the Association for Molecular Pathology (AMP) published guidelines to standardize the interpretation and classification of sequence variants in genes associated with Mendelian diseases (Richards et al., 2015). The guidelines have since been widely adopted by the genetics community, with a recent study showing that 97% of surveyed laboratories reported using the evidence criteria (Niehaus et al., 2019).

The ACMG/AMP guidelines were intended to be broadly applicable and were designed to provide flexibility and adaptability by applying expert judgment. However, recent studies have reported discrepancies and inconsistency in variant interpretation and classification between laboratories using the ACMG/AMP guidelines (Amendola et al., 2016; Amendola et al., 2020; Harrison et al., 2017). This highlights the need to develop further expert-led guidance specific to the gene or disease assessed.

The Clinical Genome Resource (ClinGen) is a National Institute of Health (NIH)-funded initiative (www.clinicalgenome.com) that aims to define the clinical relevance of genes and variants through collaborative international efforts to improve genetic diagnosis. ClinGen consists of several working groups of experts from different clinical domains. Specifically, the Variant Curation Expert Panels (VCEP) are tasked with developing gene-specific variant curation guidelines (Rivera-Munoz et al., 2018). The Glaucoma VCEP is part of the Ocular Clinical Domain Working Group. It was formed in 2019 to review the evidence and curate variants in genes associated with primary glaucoma.

Glaucoma refers to a heterogeneous group of disorders characterized by progressive optic neuropathy with associated visual field defects (Casson et al., 2012). Glaucoma is the leading cause of irreversible blindness globally. In the 2019 World Report on Vision, the WHO estimated that 6.9 million people globally have moderate or severe blindness due to glaucoma that was likely preventable (World Health Organization, 2019). Elevated intraocular pressure (IOP) is an important risk factor for glaucoma (Gordon et al., 2002; Leske et al., 1995) and evidence-based treatment consists of IOP-lowering therapies (eye drops, laser, surgery). There is clear evidence that interventions for glaucoma minimize preventable vision loss when implemented early (AGIS Investigators, 2000; Kass et al., 2002). Primary open-angle glaucoma (POAG, MIM 137750) is the most common type of glaucoma and is characterized by an open anterior chamber angle and no developmental defects or other underlying disease (American Academy of Ophthalmology Preferred Practice Pattern Glaucoma Committee, 2020). POAG with an early age of onset is called juvenile open-angle glaucoma (JOAG) and is usually defined as an age at diagnosis <30 to 40 years (but after age 5) (Turalba & Chen, 2008). POAG affects 50 to 60 million individuals worldwide, including 3% of individuals over the age of 40 years (Quigley & Broman, 2006; Tham et al., 2014).

Pathogenic variants in the *myocilin* gene (*MYOC*, formerly *trabecular meshwork-induced glucocorticoid response gene* or *TIGR*, MIM 601652) are the most common cause of Mendelian JOAG and POAG. First reported in 1997 (Stone et al., 1997), variants in the *MYOC* gene are now known to account for 8-36% of JOAG (Shimizu et al., 2000; Vincent et al., 2002; Wiggs et al., 1998) and 2–4% of POAG (Fingert et al., 1999; Souzeau et al., 2013). In high-risk individuals with a strong family history of glaucoma, cascade genetic screening can allow targeted clinical screening and help to facilitate early diagnosis and treatment of glaucoma (Souzeau et al., 2017). The *MYOC* database compiles all published variants and currently contains 336 variants (www.myocilin.com, accessed 2 September 2021) (Hewitt et al., 2008). Glaucoma causing *MYOC* variants are transmitted in an autosomal dominant manner, often with an age-related, incomplete penetrance. The most common pathogenic *MYOC* variant is p.Gln368Ter, accounting for 1.6–2.6% of individuals with POAG (Fingert et al., 1999; Souzeau et al., 2013). This variant is present in as many as 1/450 individuals in reference databases (gnomad.broadinstitute.org, version 2.1.1 accessed 2 September 2021), and previous studies support a common European founder effect (Baird et al., 2003; Fingert et al., 1999).

*MYOC* contains 3 exons and encodes a 504 amino acid protein. Most reported disease-causing variants are located in the olfactomedin domain coded by exon 3 (amino acid residues 246-502) (Hewitt et al., 2008). Both truncating and missense variants are associated with the disease. *MYOC* whole gene deletions are not associated with disease in humans (Wiggs & Vollrath, 2001) and absence of disease with heterozygous or homozygous premature termination variants in humans (Lam et al., 2000) or mouse models (Kim et al., 2001), strongly argues against haploinsufficiency as a disease mechanism. The mechanism by which *MYOC* variants cause POAG is not fully understood but current evidence supports a toxic gain-of-function mechanism. MYOC can be N-glycosylated in the endoplasmic reticulum and is efficiently secreted extracellularly (Caballero & Borrás, 2001; Caballero et al., 2000). *In vitro* studies have shown that mutant and wild-type MYOC proteins interact together to form hetero-oligomers (Gobeil et al., 2004). Evidence suggests that *MYOC* variants lead to misfolded protein (Burns et al., 2011; Liu & Vollrath, 2004; Vollrath & Liu, 2006) and that the mutant protein is not secreted extracellularly (Caballero et al., 2000; Gobeil et al., 2004; Izumi et al., 2003; Jacobson et al., 2001; Nakahara & Hulleman, 2021; Vollrath & Liu, 2006; Zadoo et al., 2016). Moreover, mutant MYOC protein is not folded correctly in the endoplasmic reticulum leading to accumulation of insoluble aggregates inside the trabecular meshwork cells (Caballero & Borrás, 2001; Liu & Vollrath, 2004), inducing the unfolded protein response and resulting in endoplasmic reticulum stress-induced apoptosis (Yam et al., 2007).

The Glaucoma VCEP was created with the goal of generating specifications to the ACMG/AMP guidelines for *MYOC*. Herein, we describe the process of developing specified variant classification rules for *MYOC*, validating the rules on a set of variants, and curating *MYOC* variants with submission to ClinVar with expert panel review status. ClinVar is a freely accessible, public archive of reports of the relationships among human variations and phenotypes, with supporting evidence (Landrum et al., 2018).

## MATERIALS AND METHODS

### VCEP membership and framework

The Glaucoma VCEP sits within the ClinGen Ocular Clinical Domain Working Group and the processes used fit within the framework provided by ClinGen for VCEPs (protocol version 9). Members were selected to provide broad expertise including clinical ophthalmology and research, with an emphasis on glaucoma as a sub-specialty, molecular biology, molecular diagnostics and genetic counselling. Representation from multiple countries was also taken into consideration, with panel members from Australia, the United States and Germany.<u>Details of composition and membership can be found</u> on the VCEP website (https://clinicalgenome.org/affiliation/50053/).

### Project design

The Glaucoma VCEP met regularly through video and telephone conferences to review the ACMG/AMP guidelines (Richards et al., 2015) and discuss changes and specifications relevant to *MYOC* POAG/JOAG. Individuals or small working groups were convened to gather relevant evidence and information for each rule, and this was presented to the VCEP for discussion. Consensus decisions were reached on the conference call and minutes circulated with the accompanying information to members unable to attend. All members were able to bring any criterion back for further discussion at any time.

Draft rules were applied to a list of 81 pilot variants, which included the majority of variants with previous classifications from ClinVar, to assess how the refined criteria performed against previous classifications. The list covered variants from all 3 exons, different categories of variants (truncating variants [nonsense, frameshift], missense, synonymous, in-frame indels), different variant classifications (benign [B]/likely benign [LB], pathogenic [P]/likely pathogenic [LP], and variant of uncertain significance [VUS]) and all variants with conflicting evidence from ClinVar). Additional variants were included to ensure that all criteria were applied in the pilot. All variants with functional evidence were included to review the level of strength of these studies. Two biocurators applied the draft rules to each pilot variant (P.G. and J.H.), noting issues that arose while doing so. These were discussed by a core pilot working group consisting of a genetic counsellor (E.S.), an ophthalmologist (D.A.M.), a molecular geneticist (K.P.B.), a clinical scientist (A.D.), and the biocurators. This group made recommendations for further modifications and refinements to the rules for discussion by the full VCEP in an iterative process. The final set of rules was disseminated to all VCEP members for comment before submission to the ClinGen Sequence Variant Interpretation Working Group, which provides harmonization across Clinical Domain Working Groups.

The ClinGen Variant Curation Interface was used to gather evidence, apply criteria, and interpret variants (https://curation.clinicalgenome.org/). Variant curations performed by the biocurators were reviewed by a core approval member (E.S.) for initial assessment, with additional review by two more core approval members (A.D., K.P.B., F.P., K.W.) for final approval. A summary of approved variant interpretations was then sent to all VCEP members for feedback. All variant classifications, criteria applied and supporting evidence were submitted to ClinVar with expert status (3-star level). Variants were annotated using GenBank reference sequence NM_000261.2 and NP_000252.1 using genome build GRCh37/hg19.

## Data sources

Publicly available variant data was obtained between June and November 2021 from ClinVar (https://www.ncbi.nlm.nih.gov/clinvar/intro/) and the Myocilin database (www.myocilin.com) (Hewitt et al.), which compiles all variants reported in the literature. Additional published and unpublished case-level data were used for variant curation from research databases of VCEP panel members including J.E.C. (The Australian and New Zealand Registry of Advanced Glaucoma [ANZRAG]) (Souzeau et al., 2013), D.A.M. (The Glaucoma Inheritance Study of Tasmania [GIST]) (Mackey et al., 2019), F.P., and T.Y. All variants are publicly listed in the Myocilin database. We defined JOAG as a diagnosis at ≤40 years old.

### Rule specific approaches

#### Population data (BA1, BS1, PM2)

Inheritance pattern, disease prevalence, allelic contribution and penetrance information were gathered from the literature and used in the Whiffin/Ware calculator (https://cardiodb.org/allelefrequencyapp/) (Whiffin et al., 2017) to establish minor allele frequency thresholds for BA1, BS1 and PM2 specific to *MYOC*. The Whiffin/Ware calculator determines a maximum credible population allele frequency based on parameters specific to the genetic architecture of the phenotype assessed. These parameters include the inheritance, disease prevalence, allelic contribution, and penetrance.

#### Proband counts (PS4)

To set thresholds for the number of probands to indicate a significant increase in prevalence of a variant in affected individuals compared to controls, odds ratios were calculated for hypothetical variants with varied numbers of observed probands and minor allele counts in population databases, similar to the approach taken by Kelly et al. (2018). Sample size was set to 3,522 cases, (based on the number of POAG/JOAG probands with *MYOC* results in the ANZRAG/GIST databases in March 2021) and 125,748 unrelated reference samples (as recorded in gnomAD v2.1.1 at the same date). Given that ANZRAG predominantly contains participants of European descent, the process was repeated with 3,277 cases and 56,885 unrelated reference samples, representing the Non-Finnish European (NFE) cohort in gnomAD. Proband count for Supporting, Moderate and Strong thresholds were selected as those that gave rise to Odds Ratios of 10, 30 and 100, as per Kelly et al. (2018).

#### Segregation data (PP1)

Thresholds for the number of meioses required to apply PP1 at each level of strength were taken from Kelly et al. (2018) and Jarvik and Browning (2016), and the classification outcomes compared. The thresholds giving the most conservative classification were selected.

#### Computational predictive tools (PP3, BP4, BP7)

Rare Exome Variant Ensemble Learner (REVEL) (Ioannidis et al., 2016), Functional Analysis Through Hidden Markov Models (FATHMM) (Shihab et al., 2013) and Combined Annotation-Dependent Depletion (CADD) (Kircher et al., 2014; Rentzsch et al., 2019) scores were calculated for all non-synonymous and synonymous (where possible) *MYOC* variants reported in ClinVar (July 2021) and compared to each other and against the classifications reported in ClinVar. HSF (Desmet et al., 2009), MaxEntScan (Yeo & Burge, 2004), NeuralNetwork (Reese et al., 1997) and SpliceAI (Jaganathan et al., 2019) were used to predict effects on splicing for synonymous variants. Tools and threshold selection were made based on the ability of each tool to discriminate variants previously classified in ClinVar as P/LP or B/LB, combined with the recommendations for interpreting output from each tool.

## RESULTS

### Summary of rule specifications

The final rule specifications adapted from the ACMG/AMP guidelines for *MYOC* by the Glaucoma VCEP were approved by the Sequence Variant Interpretation (SVI) Working Group on 12 October 2021. These are summarized in Table 1 and are available online (https://www.clinicalgenome.org/affiliation/50053/). Of the 28 criteria from the ACMG/AMP guidelines, 11 were deemed not applicable to *MYOC* (PVS1, PM1, PM3, PP2, PP4, BS2, BS4, BP1, BP2, BP3, BP5) and 2 were excluded based on previous SVI recommendations (PP5, BP6) (Biesecker & Harrison, 2018). The remaining 15 rules were all modified to be specific to *MYOC* and/or the associated phenotype. In addition, the level of strength was modified for two rules (PM2, BS3) and a scale for the level of strength was applied to nine rules (PS1, PS2, PS3, PS4, PM4, PM5, PM6, PP1, BS3). The different modifications to the rules are detailed below.

**Table 1:** *MYOC* rule specifications. JOAG: juvenile open-angle glaucoma, POAG: primary open-angle glaucoma, B: benign, LB: likely benign; LP: likely pathogenic, P: pathogenic, AA: amino acid, SVI: ClinGen Sequence Variant Interpretation Working Group

| PATHOGENIC CRITERIA | ACMG criteria | STRONG | MODERATE | SUPPORTING | Specification |
| --- | --- | --- | --- | --- | --- |
| Population data |  |  |  |  |  |
| PM2 | Absent in population databases | | | Allele frequency $\leq 0.0001$ in population databases. | Highest allele frequency in population databases should be used. Only applies to populations with $\geq 10,000$ alleles. |
| PS4 | The prevalence of the variant in affected individuals is significantly increased compared with the prevalence in controls | $\geq 15$ probands from multiple independent studies | $\geq 6$ probands from multiple independent studies | $\geq 2$ probands from multiple independent studies | Probands counted should be clinically assessed and have a diagnosis of JOAG or POAG. PM2_Supporting must be met. |
| Computational and predictive data |  |  |  |  |  |
| PP3 | Multiple lines of computational evidence support a deleterious effect on the gene or gene product | | | Applies to missense variants with a REVEL score of $\geq 0.7$. | |
| PM5 | Novel missense change at an amino acid residue where a different missense change determined to be pathogenic has been seen before | | Same residue as a previously established P variant (assessed independently of PM5) or 2 previously established LP variants (both assessed independently of PM5). | Same residue as a previously established LP variant (assessed independently of PM5). | The novel change must not affect splicing (SpliceAI $\leq 0.2$), must meet PP3 and have a Grantham score equal or greater than the previously established P/LP variants. |
| PS1 | Same amino acid change as a previously established pathogenic variant regardless of nucleotide change | Same amino acid change as a previously established P variant (assessed independently of PS1). | Same amino acid change as a previously established LP variant (assessed independently of PS1). | | The novel change must not affect splicing (SpliceAI $\leq 0.2$). |
| PM4 | Protein length changes as a result of in-frame deletions/insertions in a nonrepeat region or stop-loss variants | | In-frame del/ins and truncating variants involving $> 10\%$ of the protein. | In-frame del/ins and truncating variants involving $\leq 10\%$ of the protein. | Variant must be located within the conserved olfactomedin domain (AA 246-502). |

| Functional Data |  |  |  |  |  |
| --- | --- | --- | --- | --- | --- |
| PS3 | Well-established in vitro or in vivo, functional studies supportive of a damaging effect on the gene or gene product | Assays with OddsPath > 18.7 | Assays with OddsPath > 4.3 | Assays with OddsPath > 2.1 | Applies to the following functional studies: Solubility or secretion assays or animal models that replicate the glaucoma phenotype. |
| Segregation Data |  |  |  |  |  |
| PP1 | Cosegregation with disease in multiple affected family members in a gene definitively known to cause the disease | ≥7 meioses in >1 family | ≥ 5 meioses | ≥ 3 meioses | BA1 and BS1 must not be met. Only genotype positive/phenotype positive and obligate carriers/phenotype positive individuals should be counted as segregations. Phenotype positive need to be clinically assessed and either have a diagnosis of glaucoma or suspicious signs of glaucoma. |

| De novo Data |  |  |  |  |  |
| --- | --- | --- | --- | --- | --- |
| PS2/PM6 | De novo in a patient with disease and no family history | ≥ 2 confirmed de novo in JOAG | ≥ 2 confirmed de novo in POAG or 1 confirmed de novo in JOAG or ≥2 assumed de novo in JOAG. | 1 confirmed de novo in POAG or ≥ 2 assumed de novo in POAG or 1 assumed de novo in JOAG | Use proposed SVI point recommendations for “phenotype consistent with the gene but not highly specific and with high genetic heterogeneity” for POAG and “phenotype consistent with gene but not highly specific” for JOAG. Both maternity and paternity need to be proven for confirmed de novo variants. Parents need to be clinically assessed and not have a diagnosis of glaucoma. |

| BENIGN CRITERIA | ACMG criteria | STAND ALONE | STRONG | MODERATE | SUPPORTING | Specification |
| --- | --- | --- | --- | --- | --- | --- |
| Population data |  |  |  |  |  |  |
| BA1/BS1 | Allele frequency is greater than expected for disorder | BA1 Allele frequency ≥ 0.01 in population databases. | BS1 Allele frequency ≥ 0.001 in population databases. Does not apply to p.Gln368Ter. |  |  | The highest allele frequency in population databases should be used. Variant must be present in ≥ 5 alleles in any validated general continental population dataset of at least 2,000 observed alleles. |

| Computational and predictive data |  |  |  |  |  |  |
| --- | --- | --- | --- | --- | --- | --- |
| BP4 | Multiple lines of computational evidence suggest no impact on gene/gene product | | | | Applies to missense variants with a REVEL score $\leq 0.15$ or synonymous or noncoding variants with CADD $\leq 10$ AND SpliceAI $\leq 0.2$ | |
| BP7 | A synonymous (silent) variant for which splicing prediction algorithms predict no impact to the splice consensus sequence nor the creation of a new splice site AND the nucleotide is not highly conserved. | | | | Applies to synonymous variants or noncoding variants with no impact on splicing (SpliceAI $\leq 0.2$) AND GERP $< 0$ for conservation | |

| Functional Data |  |  |  |  |  |  |
| --- | --- | --- | --- | --- | --- | --- |
| BS3 | Well-established in vitro or in vivo functional studies show no damaging effect on protein function or splicing |  |  | Assays with OddsPath < 0.23 | Assays with OddsPath < 0.48 | Applies to variants showing solubility or secretion in functional assays. |

### Population data

#### Allele frequency data (BA1, BS1, PM2_Supporting)

BA1, BS1 and PM2 apply to the frequency of a variant in a control or general population. The Glaucoma VCEP used the Whiffin/Ware calculator to calculate allele frequency thresholds for BA1 (allele frequency >5%) and BS1 (allele frequency greater than expected for the disorder) specific to *MYOC*.

Conservative estimates from the literature were used for the disease prevalence, allelic contribution and penetrance. *MYOC* variants are associated both with POAG and JOAG. POAG is the most common phenotype, with a worldwide prevalence in 2020 estimated at approximately one in 50 individuals over the age of 40 years (Quigley & Broman, 2006; Tham et al., 2014). The highest prevalence for POAG is found in African populations (1/24) (Quigley & Broman, 2006; Tham et al., 2014). The maximum proportion of individuals with JOAG or POAG potentially attributable to a single allele corresponds to the *MYOC* p.Gln368Ter variant, which is the most frequent reported variant. Fingert et al. (1999) reported a prevalence for p.Gln368Ter of 1.6% among 1,703 individuals with POAG. Another study analyzing data from ANZRAG and GIST reported a similar prevalence of 2.6% among 1,060 individuals with POAG (Souzeau et al., 2013), with more recent data from these cohorts containing additional individuals confirming this figure (Siggs et al., 2021). Analysis of the penetrance of p.Gln368Ter showed that it was much lower in a population-based study (UK Biobank [UKB], 7.6%) than in family-based studies (ANZRAG/GIST, 56%) (Han et al., 2019), although both studies have their own unique ascertainment biases. The penetrance in the UKB was likely underestimated, given that glaucoma diagnosis relied on self-report, and it is well established that at least 50% of individuals with glaucoma are undiagnosed (Mitchell et al., 1996; Soh et al., 2021). More recent analyses indicate the penetrance of the p.Gln368Ter variant may be higher in UKB, at around 25% (Zebardast et al., 2021). Conversely, the penetrance in family-based studies may be inflated if individuals with glaucoma are more likely to participate while unaffected family members remain untested. Using the prevalence of POAG in Africans (the highest among all continental ancestries), a maximum allelic contribution at 2.6% and a conservative estimate for the penetrance at 7.6%, we calculated a maximum credible population allele frequency for BA1 at 0.007, which was rounded up to 0.01 (1%). Similarly, using the same prevalence and maximum allelic contribution, and a more realistic estimate of the penetrance at 56%, we calculated an allele frequency threshold for BS1 at 0.001 (0.1%). We adopted the SVI recommendations that the variant be present in ≥5 alleles in any validated general continental population dataset of at least 2,000 observed alleles (Ghosh et al., 2018; Sequence Variant Interpretation Working Group, 2020), to prevent using less well-characterized populations in the assessment of BA1 and BS1.

The Glaucoma VCEP decided to apply an exception to the BS1 rule for variant p.Gln368Ter based on its penetrance and the presence of a common disease haplotype in all carriers (Baird et al., 2003). Its allele frequency was 0.0025 in the UKB (Han et al., 2019) and 0.001588 in Non-Finnish Europeans in gnomAD, with the highest allele frequency at 0.003344 in Finnish Europeans, which were all above the BS1 threshold established. However, *MYOC* p.Gln368Ter is a definitively established pathogenic variant (Fingert et al., 1999; Jacobson et al., 2001; Zhou & Vollrath, 1999), displaying variable expressivity (Craig et al., 2001) and reduced penetrance (Han et al., 2019).

PM2 was initially defined by the ACMG/AMP guidelines as the absence of the variant from population databases. Although most *MYOC* pathogenic variants are absent from large population databases, they may still be present due to POAG being a common complex disease with late onset and age-related penetrance. Such phenotypes are not actively excluded from the gnomAD database. We therefore decided to allow the presence of *MYOC* variants from population databases for PM2. The filtering allele frequency for PM2 was set one order of magnitude lower than BS1 at 0.0001 (0.01%). PM2 only applies to populations of ≥ 10,000 alleles, to prevent the occurrence of a variant by chance in a small population subset. The Glaucoma VCEP endorsed the SVI recommendation to decrease the weight of PM2 to a Supporting level (Sequence Variant Interpretation Working Group, 2020). Finally, we recommended using the highest allele frequency in population databases when assessing BA1, BS1 and PM2_Supporting.

#### Phenotype data (PS4, PS4_Moderate, PS4_Supporting)

On review of the literature, reliable case-control data in individual studies was limited for most *MYOC* variants, with the size of the control cohort often not large enough for a reliable estimate of frequency. Therefore, the Glaucoma VCEP decided to use the “proband counting” approach recommended by the ACMG/AMP guidelines, which allows counting of probands across multiple independent studies for a “quasi case-control study”.

All probands counted must be clinically assessed and have had a diagnosis of JOAG or POAG. Due to incomplete penetrance, late age of glaucoma diagnosis and the rate of undiagnosed glaucoma in the general population, pathogenic variants are expected to be present in population databases. Therefore, PM2_Supporting (frequency <0.0001 in control population) should be met (indicating a rare allele) in order to apply PS4. However, BA1 and BS1 (frequency in control populations) must not be met to avoid counting the occurrence of common variants in affected individuals. Efforts should be made to ensure that probands counted are from independent cohorts and probands should not be counted toward PS4 if uncertainty remains (e.g., published by same groups or authors).

We calculated proband count thresholds at which statistical significance would be reached when compared to a large reference dataset such as gnomAD. Evaluation of the ANZRAG/GIST cohorts as a basis for calculations revealed that no pathogenic variants from the cohorts were present at more than 6 alleles in gnomAD (excluding p.Gln368Ter). Using the presence of 6 alleles in gnomAD, ORs of 10, 30 and 100, as per Kelly et al. (2018), were reached for 2, 6 and 17 probands (Supplementary Table 1). Calculations on cases and controls from a European-only population gave similar thresholds of 2, 6 and 18 probands for ORs of 10, 30 and 100 respectively, when 3 alleles were present in the reduced gnomAD dataset (Supplementary Table 1). Based on results similar to Kelly et al., the same thresholds of 2, 6, and 15 probands for Supporting, Moderate and Strong were selected for the application of PS4.

### Computational and predictive data

#### Computational predictive tools (PP3, BP4, BP7)

Evaluation of a broad range of computational predictive tools revealed considerable overlap in the evidence used to make predictions. The most commonly used tools (CADD, REVEL, FATHMM) aggregate information from multiple features. In order not to overstate the importance of computational evidence, and in line with reports that a lower rate of concordance is achieved when multiple software tools are used (Ghosh et al., 2017), the Glaucoma VCEP decided to use a single tool that collates multiple sources of computational evidence in the first instance, and where necessary, supplement that with additional sources of evidence.

A comparison of REVEL, FATHMM and CADD was undertaken for nonsynonymous variants deposited in ClinVar. FATHMM appeared to be optimized towards assessing non-coding variants, which are of little relevance in *MYOC* glaucoma and were thus discounted.Comparison of the REVEL and CADD scores revealed that REVEL was more conservative (Supplementary Figure 1). CADD inappropriately scored multiple benign variants in exon 1 as pathogenic, whereas REVEL did not. Multiple benign and VUS variants had CADD scores >20, but REVEL scores <0.5 However, there was broad agreement between the tools at the pathogenic end.. Based on this comparison, and the sensitivity/specificity reported for REVEL when predicting pathogenicity of ClinVar variants (Ioannidis et al., 2016), the Glaucoma VCEP recommended defining PP3 as REVEL ≥0.7 (58% sensitivity, 96% specificity) and BP4 as REVEL ≤0.15 (55% sensitivity, 95% specificity) for missense variants. This specification is in line with other VCEPs curating variants for autosomal dominant diseases (Luo et al., 2019; Oza et al., 2018). The CADD scores for truncating variants did not correlate well with Clinvar classifications, with five truncating variants assessed scoring >30, but only two were clearly pathogenic. REVEL does not score truncating variants. It was decided that truncating variants should be considered under PM4 (protein length changing variants), given the uncertainties around *in silico* prediction of the effect of truncating MYOC.

Several criteria require specific evaluation of potential splicing effects, which may apply to missense variants but equally to synonymous or non-coding variants. While REVEL takes into account splicing effects for missense variants, it is unable to score synonymous variants. We evaluated several splice prediction tools for this purpose, seeking one that was freely available as well as straightforward to use and interpret the output. SpliceAI (Jaganathan et al., 2019) met these criteria and had clear interpretation guides in the reporting literature. We did not find any evidence in the literature of *MYOC* variants (synonymous or non-synonymous) believed to act through altered splicing and all 19 synonymous variants reported in ClinVar at the time of assessment were classified as B/LB, VUS or conflicting interpretations. All of these had SpliceAI scores <0.2 for all four assessed splicing measures, consistent with no *in silico* evidence of functional effects on splicing. Therefore, we chose a SpliceAI scores ≤ 0.2 for assessing splicing effects of synonymous variants.

BP4 requires multiple lines of computational evidence suggesting no impact on gene product. As CADD scores can be calculated for any type of variant and incorporate multiple lines of evidence, we specified that a CADD score of ≤10 and SpliceAI score ≤0.2 were required to apply BP4 to synonymous variants. SpliceAI is not incorporated into the CADD model, and the addition of this algorithm gives added confidence towards a benign classification when the two tools agree. For the application of BP7 for synonymous variants, we specified a GERP score <0, indicating the nucleotide is not highly conserved, as well as SpliceAI ≤0.2.

The Glaucoma VCEP agreed to extend BP4 and BP7 to noncoding variants. However, based on the disease mechanism and the absence of current evidence supporting pathogenicity of intronic/noncoding variants, we decided that the curation of these variants would be deprioritized.

#### Protein length changing variants (PM4, PM4_Supporting)

The Glaucoma VCEP decided that PM4 (protein length changes due to in-frame deletions/insertions and stop losses) would be more appropriate for truncating variants in the last exon that are predicted to escape nonsense-mediated decay, instead of PVS1, which is intended for loss-of-function (LoF) impact. PM4 applies to in-frame deletions or insertions and stop-loss variants as per the ACMG/AMP guidelines. We decided to expand PM4 to any protein length-changing variants, including truncating variants, located within the conserved olfactomedin domain. The ACMG/AMP guidelines emphasized that larger deletions, insertions or extensions would be considered stronger evidence toward pathogenicity.

Therefore, we decided to apply PM4 at a Moderate level if the protein length-changing variant was involving ≥10% of the protein and at a Supporting level if involving <10% of the protein.

#### Variants affecting the same amino acid residue (PS1, PS1_Moderate, PM5, PM5_Supporting)

The ACMG/AMP guidelines defined two criteria affecting the same amino acid residue as a previously established pathogenic variant: PS1 applies to a novel nucleotide change leading to the same amino acid residue while PM5 applies to a different missense change. There was only one variant listed in the *MYOC* database that consists of a different nucleotide leading to the same amino acid (p.Asn480Lys caused by c.1440C>A and c.1440C>G). In contrast, there were 24 variants in the *MYOC* database with 2 different missense variants reported at the same residue and 6 with >2 different missense variants reported at the same residue. The Glaucoma VCEP decided to apply different levels of strength to PS1 and PM5 depending on the pathogenicity of the previously established variant. PS1 applies to the same amino acid change as a previously established P variant while PS1_Moderate applies to the same amino acid change as a previously established LP variant. The previously established P or LP variants need to reach their classification without the use of PS1.

Similarly, PM5 applies to the same residue as a previously established P variant or to two previously established LP variants while PM5_Supporting applies to the same residue as a previously established LP variant. The previously established P or LP variants need to reach their classification without the use of PM5. The ACMG/AMP guidelines advised caution for changes that may impact splicing rather than act at the amino acid/protein level. Therefore, we specified that the novel change must not affect splicing for PS1 and PM5 to apply (as assessed by SpliceAI with a score ≤ 0.2). Additionally, the missense variant assessed must meet PP3 and have a Grantham score equal or greater than the previously established P or LP variant to ensure the novel amino acid is predicted to impact function to apply PM5.

### Functional data (PS3, PS3_Moderate, PS3_Supporting, BS3_Moderate, BS3_Supporting)

PS3 applies to functional evidence from well-established *in vitro* or *in vivo* functional studies supportive of a damaging effect on the gene or gene product. The current body of literature supports a role for *MYOC* variants causing JOAG or POAG via a mechanism by which the mutant protein is not secreted extracellularly (Caballero et al., 2000; Gobeil et al., 2004; Izumi et al., 2003; Jacobson et al., 2001; Vollrath & Liu, 2006), which leads to the accumulation of insoluble aggregates in the cells (Caballero & Borrás, 2001; Liu & Vollrath, 2004). The Glaucoma VCEP reviewed the different assays used to assess MYOC function. We determined that assays that report on the solubility and secretion of MYOC were considered suitable, with functional evidence supporting insolubility and non-secretion of mutant MYOC protein indicating impact on protein function. Transgenic mice expressing the Tyr423His MYOC mutant (corresponding to human MYOC p.Tyr437His) (Senatorov et al., 2006), or the human p.Tyr437His (Zode et al., 2011), develop elevated IOP, retinal ganglion cell death and axonal degeneration, reproducing the glaucoma phenotype seen in humans. We determined that animal models that replicate the glaucoma phenotype were also suitable.

We decided to follow the recommendations from Brnich et al. (2019) that PS3 should only be applied if the assay includes both negative (e.g., empty vector) and positive (e.g., wild-type) controls and includes technical and/or biological replicates. We decided to apply different levels of strength based on the recommended odds of pathogenicity (OddsPath) (Brnich et al., 2019). PS3 can be applied at a Strong, Moderate or Supporting level for OddsPath of >18.7, >4.3 and >2.1 respectively. The OddsPath can be determined based on the number of validation controls included in a particular assay, which are variants classified as P/LP (pathogenic control) or B/LB (benign control) without the use of PS3 or BS3.

We also reviewed the evidence on the solubility and secretion assays for benign variants. All coding variants curated in the pilot phase as B/LB for which functional evidence was available were secreted and/or soluble. Similarly, among the coding variants showing evidence for solubility and/or secretion, none were classified as P/LP. Therefore, we decided to apply BS3 to variants showing solubility or secretion in functional assays that meet the OddsPath of <0.48 or <0.23 for a Supporting or Moderate level, respectively (Brnich et al., 2019). Although there is no evidence currently supporting other protein functions leading to the condition, we recommended not applying BS3 at a Strong level, as we could not completely rule out other mechanisms for pathogenicity.

The Glaucoma VCEP reviewed all variants included in functional assays that reported on the solubility or the secretion of MYOC (Supplementary Tables 2a and 2b). Controls from studies that reported on the same class of assay and had the same methodology were combined to calculate the OddsPath. The number of pathogenic and benign validation controls from each study determined if PS3 and BS3 were applied and if so, at which level of strength.

We recommended that if multiple results from functional assays are available for a single variant, then the evidence from the assay that is best validated should apply as suggested (Brnich et al., 2019). If results from different assays are conflicting for a single variant, then the level of validation of each assay should be considered and the result from the assay with the highest level of validation and a conclusive result could override the result from the other assay. However, if the results from the study with the highest level of validation are inconclusive (partial secretion or insoluble levels), then PS3/BS3 should not be applied.

### Segregation data (PP1_Strong, PP1_Moderate, PP1)

In the ACMG/AMP guidelines, PP1 applies for co-segregation with disease but is not quantitatively defined. Two studies have since provided quantitative guidelines for assessing segregation of a variant and applying PP1 at different strength levels. Jarvik et al. calculated a probability that the observed variant and affection status occurs by chance rather than due to co-segregation (Jarvik & Browning, 2016). Under a dominant model, this probability is N = (1/2)m, where m is the number of meioses of the variant of interest. Similarly, Kelly et al. (2018) adopted different numbers of meioses based on likelihood ratios of 10 (LOD 0.9), 30 (LOD 1.5) and 100 (LOD 2.1) for Supporting, Moderate and Strong evidence. The pilot study compared the thresholds for the number of meioses required to apply each level of evidence described by Kelly et al. (2018) and Jarvik and Browning (2016). Among the 81 variants curated in the pilot phase, only two had a different strength of evidence based on the two approaches: p.Ala363Thr and p.Asn480Lys. The p.Ala363Thr variant had 5 meioses in 3 families, meeting PP1_Moderate according to Kelly et al. but PP1_Strong according to Jarvik and Browning, but both approaches resulted in an LP classification. The p.Asn480Lys (c.1440C>G) variant had 4 meioses in 1 family, which met PP1 according to Kelly et al. but PP1_Moderate according to Jarvik and Browning. The variant was classified as LP when using Kelly et al. but as P when using Jarvik and Browning. The Glaucoma VCEP agreed to apply a conservative approach and use the recommendations from Kelly et al. (2018) with ≥7, ≥5, and ≥3 meioses for Strong, Moderate and Supporting levels. In addition, a Strong level of evidence should only be applied if the variant is present in more than one family, to limit the risk of detecting segregation with a second variant in strong linkage disequilibrium with the one under assessment.

Benign variants may segregate in a family by chance or appear to segregate because they are common in the population. The Glaucoma VCEP agreed that PP1 would only apply if BA1 or BS1 are not met. Due to the late age of onset of POAG, the documented reduced penetrance of many variants, and the potential for phenocopies within families, only genotype-positive/phenotype-positive individuals and obligate carriers/phenotype-positive should be counted. Individuals who carry the variant but do not have a diagnosis of JOAG/POAG (genotype-positive/phenotype-negative) should not be included when counting meioses nor should JOAG/POAG patients who do not carry the variant (genotype-negative/phenotype-positive). Phenotype-positive individuals need to have been clinically assessed and either have a diagnosis of glaucoma (POAG or JOAG) or suspicious signs of glaucoma (e.g., ocular hypertension, suspicious discs).

### De novo data (PS2/PM6_Strong, PS2_Moderate/PM6, PS2_Supporting/PM6_Supporting)

PS2 and PM6 apply to *de novo* variants. *MYOC de novo* variants are rare, with only two reports in the literature: p.Val251Ala (Kuchtey et al., 2013) and p.Pro254Leu (Souzeau et al., 2016), both confirmed as *de novo*. The Glaucoma VCEP adopted the SVI-proposed point recommendation table to determine the appropriate strength of evidence for PS2 and PM6 (Supplementary Figure 2) (Sequence Variant Interpretation Working Group, 2021). The phenotype of POAG is not highly specific and has substantial genetic heterogeneity whereas the phenotype of JOAG is more specific. We recommended applying the point-based system for a “phenotype consistent with the gene but not highly specific and with high genetic heterogeneity” for POAG and a higher point-based system for JOAG, using a “phenotype consistent with gene but not highly specific”. In addition to the absence of the variant in both parents, both paternity and maternity need to be confirmed to demonstrate a variant is *de novo*, as per the ACMG/AMP guidelines. Both parents need to be clinically assessed and should not have a diagnosis of glaucoma. If a parent has suspicious signs of glaucoma, the age and the severity of the symptoms should be taken into account before applying criteria. Using these guidelines, PS2 applies to ≥2 confirmed de novo occurrences in JOAG, PS2_Moderate or PM6 to ≥2 confirmed de novo in POAG or 1 confirmed de novo in JOAG or ≥2 assumed de novo in JOAG and PS2_Supporting or PM6_Supporting to 1 confirmed de novo in POAG or ≥2 assumed de novo in POAG or 1 assumed de novo in JOAG.

### Rules removed or deemed not applicable

Two rules were removed and 11 were deemed not applicable, including 6 rules of the pathogenic framework and 7 rules of the benign framework. The reasons for each criterion are briefly explained here. PP5 (reputable source reports as pathogenic) and BP6 (reputable source reports as benign) were removed based on the recommendation from the ClinGen SVI Working Group to use primary data instead of relying on assertions (Biesecker & Harrison, 2018).

Because *MYOC* variants cause disease through a gain-of-function mechanism, PVS1 (null variant in a gene where loss of function is a known mechanism of disease) was not applicable and PM4 was considered more appropriate for truncating variants. *MYOC* has no mutational hot spot and benign variants are present throughout the well-characterized olfactomedin domain in exon 3, making PM1 (located in a mutational hot spot and/or functional domain without benign variation) not applicable. As *MYOC* is associated with an autosomal dominant condition, PM3 (detected in trans with a pathogenic variant in a recessive gene) did not apply. PP2 (missense variant in a gene with a low rate of benign missense variation) was not relevant to *MYOC*: although pathogenic missense variants are common in *MYOC*, the gene also has a significant amount of benign missense variants as shown by the missense constraint z score in gnomAD (z = 0.52), supporting tolerance to variation. PP4 (patient’s phenotype or family history highly specific for a disease with a single genetic etiology) was not applicable due to the genetic heterogeneity of POAG/JOAG and the phenotype being nonspecific.

The incomplete penetrance and the late age of onset of *MYOC*-associated glaucoma (Craig et al., 2001; Hewitt et al., 2007; Mackey et al., 2003) rendered BS2 (observed in a healthy adult individual with full penetrance expected at an early age) not applicable. Similarly, the Glaucoma VCEP decided to not use BS4 (lack of segregation in affected members of a family) due to the presence of phenocopies (Angius et al., 2000; Craig et al., 2001), reduced age-related penetrance (Craig et al., 2001; Hewitt et al., 2007; Mackey et al., 2003) and the possibility that more than one pathogenic variant can contribute to the phenotype observed in families (Hewitt et al., 2007), which make non-segregation difficult to assess. Both truncating and missense *MYOC* variants are causative, making BP1 (missense variant in a gene with primarily truncating variants) not applicable. The Glaucoma VCEP decided to not apply BP2 (observed *in trans* with a pathogenic variant for a fully penetrant dominant gene/disorder or observed *in cis* with a pathogenic variant in any inheritance pattern): biallelic *MYOC* variants (either compound heterozygotes or homozygotes) have been reported (with variable phenotype) (Wirtz et al., 2008; Young et al., 2012) and are not incompatible with life, and variants *in cis* may act synergistically while the effect of a variant occurring after a truncating variant may not be predicted. *MYOC* lacks repetitive regions of unknown function to apply BP3 (in-frame del/ins in a repetitive region without a known function). Multiple molecular diagnoses are possible and variants in different genes could have an additive effect, therefore BP5 (variant found in a case with an alternate molecular basis for disease) did not apply.

### Combining criteria

Tavtigian et al. (2018) showed that the ACMG/AMP guidelines were compatible with a Bayesian framework and further demonstrated that they could be converted into a point-based system (Tavtigian et al., 2020). This approach is especially helpful for variants with conflicting evidence that would have previously led to a VUS classification. With the point-based system, variants with conflicting evidence can be classified as pathogenic or benign depending on the number and/or the strength of the criteria applied. Additionally, this approach allows the use of criteria strength combinations not specifically listed in the ACMG/AMP guidelines. The Glaucoma VCEP decided to apply the scaled point system recently developed by Tavtigian et al. (2020) rather than the ACMG/AMP variant classification guidelines (Tables 2 and 3). We modified the threshold for LB from -1 to -2 to follow the ACMG/AMP guidelines that require ≥2 Benign Supporting criteria for a LB classification.

**Table 2:** Point Values for ACMG/AMP strength of evidence categories (Tavtigian et al., 2020)

| Evidence Strength | Point scale |  |
| --- | --- | --- |
|  | Pathogenic | Benign |
| Indeterminate | 0 | 0 |
| Supporting | 1 | -1 |
| Moderate | 2 | -2 |
| Strong | 4 | -4 |
| Very Strong | 8 | -8 |

**Table 3:** Point-based variant classification categories, modified from Tavtigian et al. (2020)

| Category | Point ranges |
| --- | --- |
| Pathogenic | $\geq 10$ |
| Likely Pathogenic | 6 to 9 |
| Uncertain | -1 to 5 |
| Likely Benign | -2 to -6 |
| Benign | $\leq -7$ |

### Application of the rules in a pilot study

We curated 81 *MYOC* variants (Supplementary Table 3) using the Glaucoma VCEP specified rules (Table 1). The majority of variants (74%, 60/81) were missense (Figure 1) and were located in exon 3 (78%, 63/81) where the largest numbers of causative variants have been reported.

**Figure 1:**
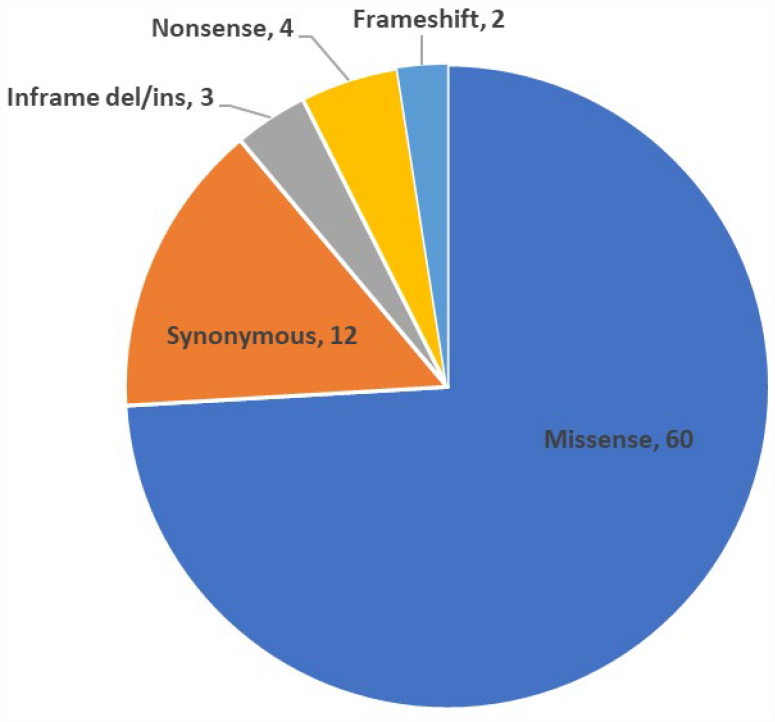
Distribution of 81 *MYOC* variants classified in the pilot study

Figure 2 shows the final classification of the pilot variants by the Glaucoma VCEP. Approximately one-third were classified as P/LP, one-third as VUS and one-third as LB/B, reflecting the intention to select roughly equal numbers of variants from each category for the pilot study. Figure 2b shows the position of the pilot variants in the *MYOC* gene. All of the LP and P variants were located in exon 3. The B, LB and VUS variants were located in each of the 3 *MYOC* exons Based on the classification results presented in Supplementary Table 3, Figure 3 shows the distribution of the classification scores for the variants, calculated by adding the points awarded for each criteria met. All of the variants classified as B met the stand alone, BA1, population criterion. No variants reached a score of ≤-7. All of the variants with a score of -6 met BS1 as well as BS3_Moderate or the combination of the computational criteria BP4 and BP7. The variants classified on the upper end of LB, with a score of -2, all solely met the functional BS3_Moderate criterion. The 25 variants classified as VUS were spread across the full classification range, from -1 to 5 points. A score of 6 was the most common for those classified as LP, with 8 variants meeting this score. Half of the P variants received a classification score of 12, which was the highest of the pilot variants.

**Figure 2:**
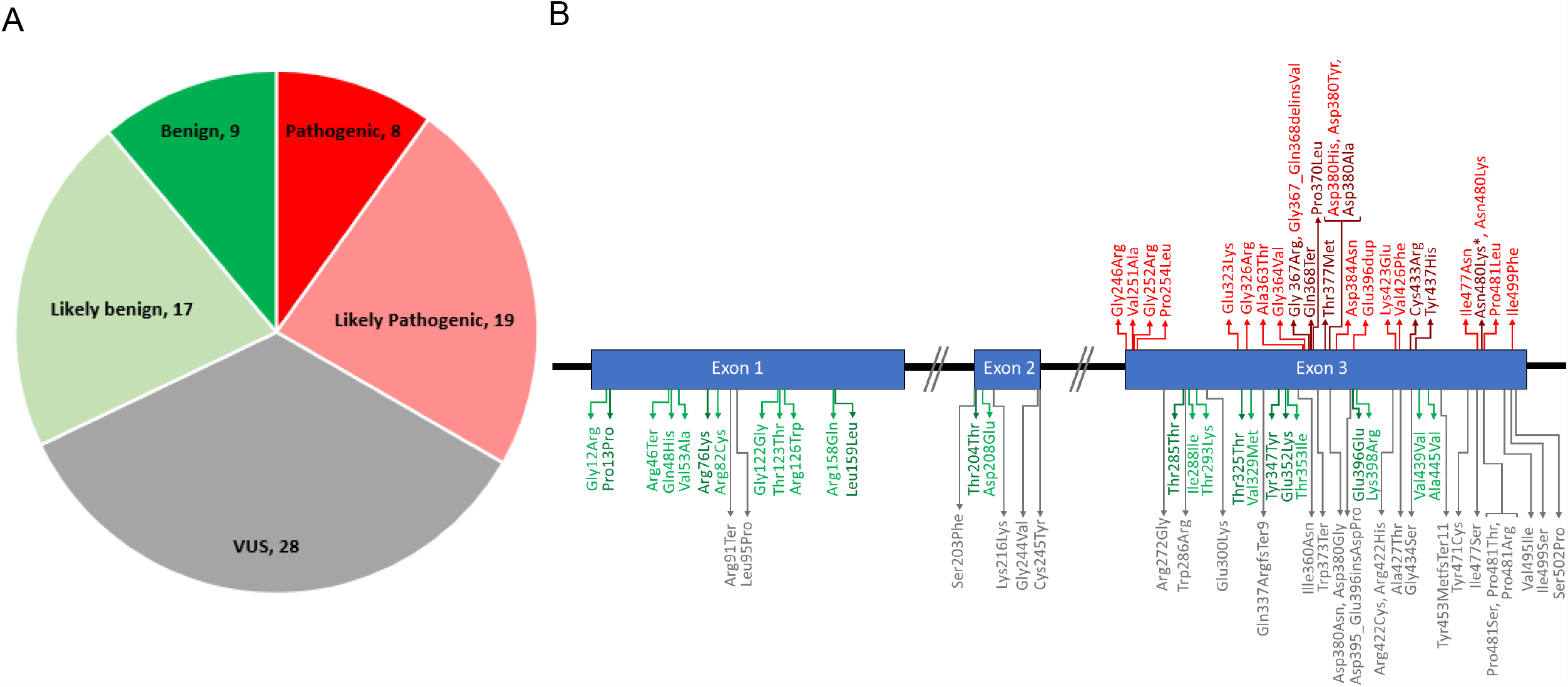
Final classification of pilot variants by the Glaucoma VCEP using the specified rules. A) number of variants in each classification. B) position of each variant in the *MYOC* gene. Dark red = P; Light red = LP; Grey = VUS; Light green = LB; Dark green = B. *there are 2 variants encoding Asn480Lys, c.1440C>A was classified as P and c.1440C>G as LP.

**Figure 3:**
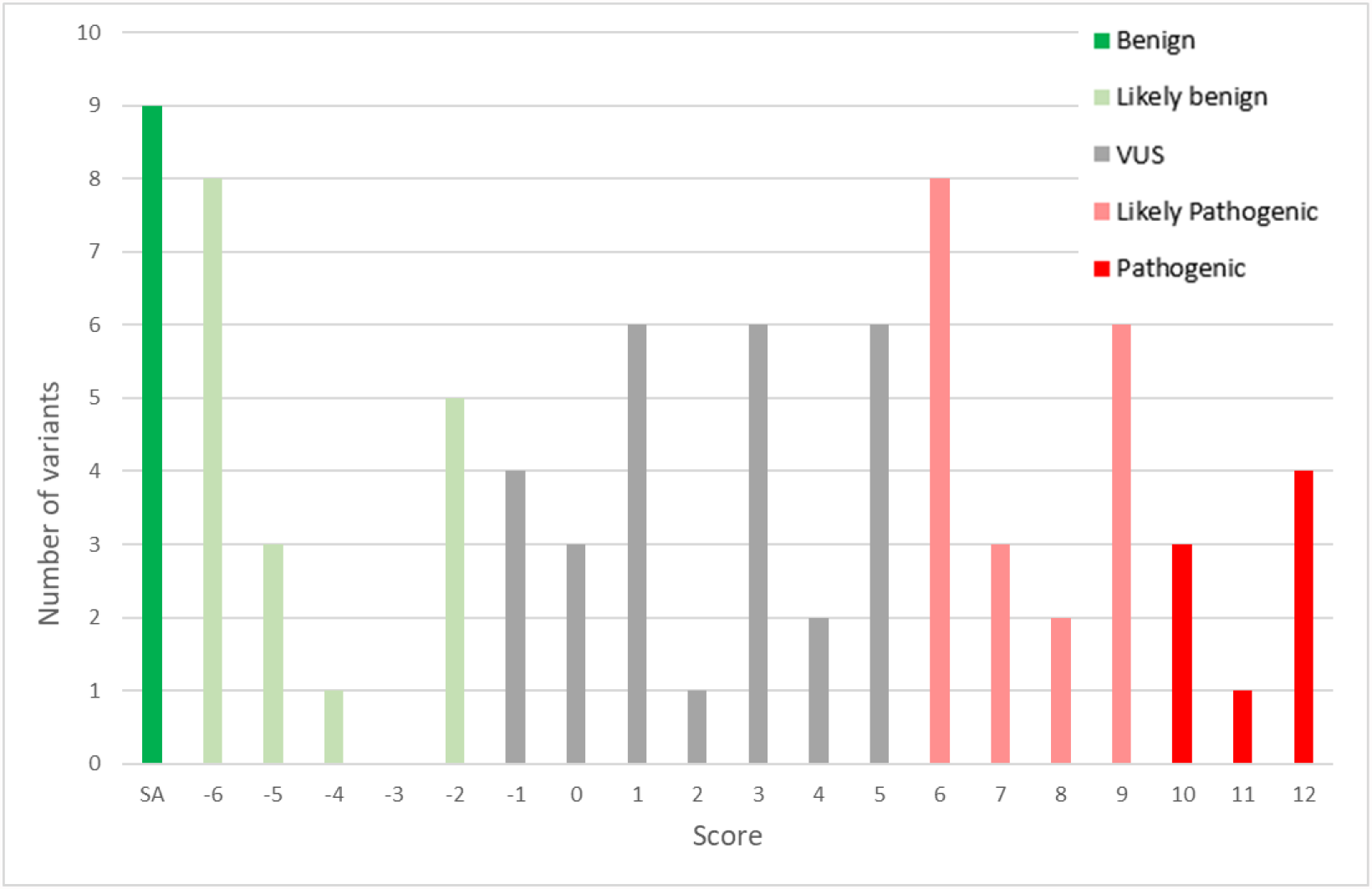
Distribution of the final Glaucoma VCEP classification scores

The stand-alone population criterion, BA1, was used to classify 9 of the pilot variants as B. Although the computational criteria BP4 and BP7, and the functional criterion, BS3_Supporting were also applied to these variants, they were not necessary for the benign classification. The population and functional criteria BS1 and BS3_Moderate were the most commonly used criteria for the variants classified as LB (applied 12 and 11 times respectively).

Five variants had conflicting evidence, with both pathogenic and benign criteria applied. The functional criterion BS3_Moderate was included in each of these classifications as was PM2_Supporting and/or PP3. All of these variants were classified as VUS, with scores not reaching the thresholds for either benign or pathogenic classifications.

Supplementary Figure 3 shows that most variants classified as LP or P had PM2_Supporting applied and were not reported in gnomAD, with the exception of the two well-characterized common variants p.Gln368Stop and p.Thr377Met. It is also evident that BS1 plays an important role in discriminating LB variants from VUS, with 81% (21/26) of variants classified as LB/B meeting BS1 or BA1.

PS1 was used once in the pilot variants, for c.1440C>G p.Asn480Lys. The Glaucoma VCEP had classified the c.1440C>A variant as pathogenic, which allowed PS1 to be applied to the other variant located at the same amino acid residue, increasing the classification score from 5 to 9 and providing this variant with a LP classification.

PM5 was used once with an unmodified strength and four times as PM5_Supporting. Based on the LP classification of p.Asp380His, PM5_Supporting was applied to p.Asp380Ala, which has a higher Grantham score, although this variant reached a P classification regardless of the application of this criterion. This pathogenic variant was then used to apply PM5 to p.Asp380Tyr, as it has a higher Grantham score than p.Asp380Ala, increasing its classification to LP. The LP variant p.Asp380His was also used to apply PM5_Supporting to p.Asp380Gly, although the final classification for this variant was VUS. Additionally, PM5_Supporting was applied to p.Pro481Arg and p.Ile499Ser, but these variants also remained as VUS.

For variants resulting in altered protein length, PM4 was applied to one frameshift and two nonsense variants while PM4_Supporting was applied to three in frame del/ins and one frameshift variant. Of these 7 variants, 4 were classified as VUS, 2 as LP and 1 as P.

The Glaucoma VCEP reviewed evidence from 10 published studies with functional assays, which included 63 variants from the pilot list. BS3_Moderate was applied 20 times in total, with 11 of these involved with LB classifications. For 5 variants, BS3_Moderate was the only criterion applied, which resulted in a score of -2 and an LB classification. PS3 was applied 31 times, 27 at the Moderate level and 4 at the Supporting level. Functional evidence influenced the classification for 29% (18/63) of variants, including 10 from VUS to LP, 5 from VUS to LB and 3 from LP to P.

Among the 40 variants that had a ClinVar classification (accessed 10 January 2022), 40% (16/40) had a change of classification after Glaucoma VCEP curation (Figure 5). Variants with a discordant classification from ClinVar included 7 VUS reclassified as B/LB, 3 LP/P reclassified (2 as VUS and 1 as B/LB) and all 6 variants with conflicting interpretation reclassified (5 as B/LB and 1 as VUS). The population criteria BA1 and BS1 were applied in the curations of 8 of the 13 variants reclassified as B/LB by the Glaucoma VCEP. BS3_Moderate was applied to 10 of these variants. The number of variants in ClinVar classified as VUS or as variants with conflicting interpretations decreased from 13 to 1 after VCEP curation.

**Figure 4:**
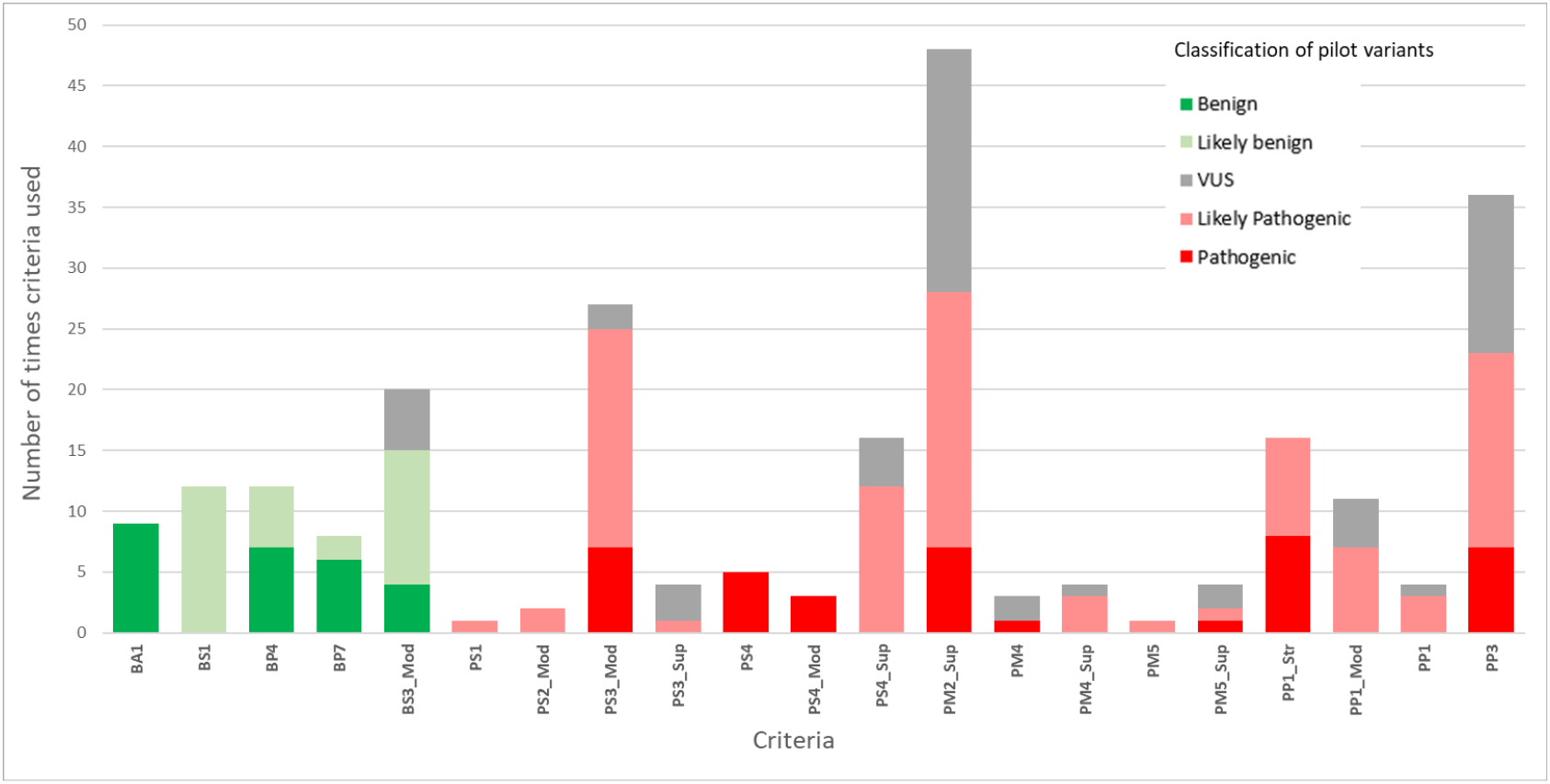
Criteria applied in the pilot phase with final Glaucoma VCEP classification of variants

**Figure 5:**
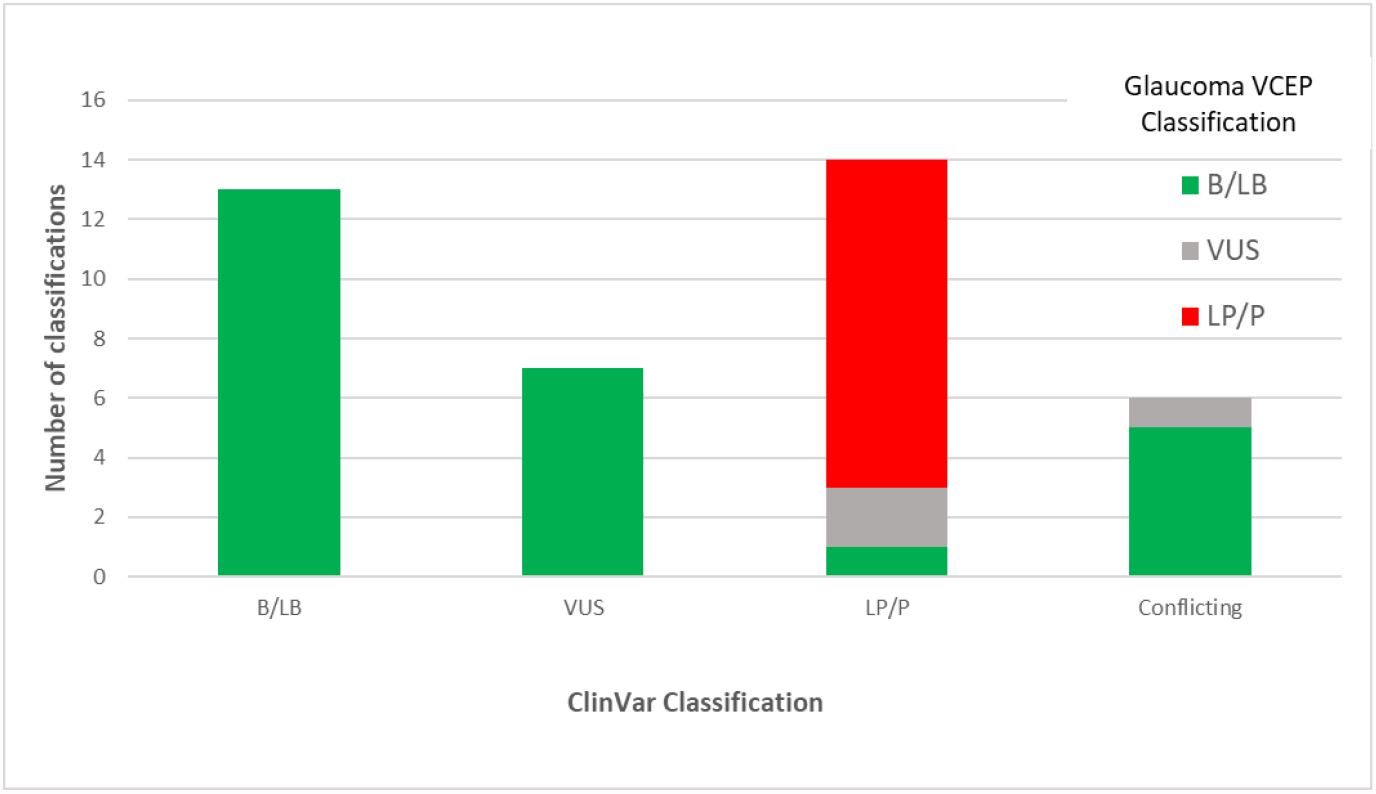
ClinVar vs Glaucoma VCEP classification of *MYOC* variants from the pilot phase. Colours indicate the Glaucoma VCEP classification

## DISCUSSION

The specification of the ACMG/AMP guidelines for the *MYOC* gene in relation to POAG/JOAG has resulted in the reclassification of over one-third of the variants reported in ClinVar. The process has led to fewer VUS, with clearer definitions of LB and B, largely related to population frequency and functional evidence. Although *in silico* predictions supported the B and LB classifications, these predictions were less critical in the final determination. Similarly, for LP and P classifications, population and functional criteria were influential, but in contrast to the benign variants, *in silico* predictions were frequently applied.

The pilot project to test the rules revealed several challenges, particularly where information from previously classified variants was required to classify a variant of interest. This was particularly evident in the application of PS1, PM5 and PS3/BS3. In these situations, we were first required to identify variants in our pilot list that met the LP or P classifications without the application of these rules before we could apply these rules to other variants. For example, PM5_Supporting or PM5 can only be applied if another variant at the same residue was previously classified as LP or P and the novel variant must have a higher Grantham score than the previously classified variant. This means the rule is not just applied at the highest level for any novel variant at the same residue as a previously reported variant and the biocurators were required to seek information about many other variants in order to apply the rule to one variant. Similarly, for PS3 and BS3, to determine the quality of the assay and the strength at which each rule can be applied, it was necessary to count the number of variants assessed by that assay that were classified B/LB or LP/P without functional evidence, and label these as validation controls for that assay. The Glaucoma VCEP expanded the pilot variant list to include all variants for which functional evaluation was identified in the literature to facilitate this process. There is unlikely to be functional evidence available for variants not included in this pilot. Generating such evidence for additional variants would be highly valuable, especially for variants classified as VUS in the absence of functional data and the literature should be regularly reviewed for such information.

The addition of carefully assessed functional data to the classification resulted in 15 variants meeting the thresholds for LB or LP that otherwise would have remained as VUS. It is tempting to apply BS3 and PS3 at high-strength levels on the basis of any published functional data. However, the Glaucoma VCEP recommends a more cautious approach given the uncertainties around the pathogenic mechanisms of glaucoma development. The work by Brnich et al. (2019) to quantify the required numbers of validation controls and confirmed pathogenic or benign variants was critical to the determination of how to apply published data, but even with the conservative evidence-based thresholds chosen, these criteria played an important role in the classification of *MYOC* variants. The VCEP chose to specify the application of BS3 at the Moderate level when the appropriate OddsPath was reached, as described by Brnich et al. This is in contrast to the original ACMG/AMP guidelines, which did not allow for any benign criteria to be applied at the Moderate level.

The application of BS3_Moderate allowed variants to be classified as LB with the application of BS3_Moderate alone (score of -2). This decision was taken to reflect a consistent approach in the level of evidence required for both BS3 and PS3 and reflects the level of confidence the Glaucoma VCEP had in the quality and validity of the functional assays described for MYOC.

Similarly, despite the rule in the original guidelines being at the Supporting level, PP1 is likely to be applied arbitrarily at higher levels. The work by Kelly et al. (2018) in relation to the *MYH7* gene and Jarvik and Browning (2016) for more general application, provide an evidence base for rational decision-making on when to apply each level of evidence. Both are simple to apply, requiring only the counting of meioses (excluding the proband) with thresholds for each level of significance correlating to the probabilities of multiple co-segregating transmissions. We chose the slightly more conservative classifications of Kelly et al. to limit over-interpretation; however, the marginal differences in outcome reflect the similarity in the underlying approaches. PS4 was originally defined for case-control studies, or, when they do not reach statistical significance, for multiple independent affected individuals. Similar to PP1, we have developed an easy-to-use points system, which allows counting of independent probands and the application of PS4 at different levels of evidence.

The benign criteria for population data BA1 and BS1 were originally defined as an allele frequency >0.05 and greater than expected for the disease, respectively. However, it has since been recognized that the prevalence and penetrance of the disease as well as the gene contribution should be considered when applying these rules (Whiffin et al., 2017). The Glaucoma VCEP has developed allele frequency thresholds for BA1 and BS1 that are more conservative and reflect the architecture of the disease and the gene assessed. This was supported by our pilot data showing that 81% of LB/B variants met one of these criteria.

Similarly, PM2 was initially defined as an absence of the variant from controls. However, we set a conservative threshold to account for the possibility of pathogenic variants being present in population databases in the context of glaucoma often being undiagnosed and having an incomplete age-related penetrance. Nevertheless, all but two of the variants classified as LP/P from the pilot study were absent from gnomAD, highlighting the rarity of pathogenic *MYOC* variants in population databases. Although the Glaucoma VCEP used gnomAD for assessing allele frequency in the pilot phase, we encourage the use of other large population datasets that become available or that may be specific to some populations.

The Glaucoma VCEP accessed allele frequency information from gnomAD and recommended applying the most relevant population specific information where available. However, it should be noted that gnomAD and most other population databases are limited in their content from non-European populations. Our pilot variant list included several variants predominantly reported in non-European probands. Several were classified as B (p.Pro13Pro seen in African and African Americans), or LB (p.Gly12Arg, p.Arg46Ter, p.Gln48His, p.Asp208Glu, p.Thr353Ile reported in East Asian/South Asian). Others were classified as VUS (p.Trp286Arg in Latino/Admixed American and p.Glu300Lys in East Asian) and two as P (p.Thr377Met and p.Cys433Arg -African/African American). As access to genetic research and testing increases in populations of non-European descent, sourcing appropriate population frequencies will be critically important for correctly interpreting novel variants. New information from other populations should also be considered for variants already classified, that may have inadvertently been labelled rare based on European information but are more common in other populations.

Access to high quality data is important for criteria requiring counting of genotype- and phenotype-positive individuals (PS4, PP1). In the pilot study, this was largely achieved through manual curation of published literature, including, where necessary, contacting the corresponding authors to confirm if individuals/families with the same variant in different publications were, in fact, the same individual reported multiple times in various contexts. Where this information could not be obtained directly, we took the conservative approach of only counting each possibly duplicated patient once. In addition, the Glaucoma VCEP accessed multiple research databases to supplement published information, increasing the number of probands counted for some variants. This highlights the importance of individual research groups and accredited genetic testing laboratories publishing and/or depositing the information for each observed variant in publicly accessible locations. Once a variant reaches an LP or P classification, further evidence is not required, but for rarely observed variants, every piece of information is useful to curators and can have a significant effect on the overall classification.

The thresholds for each criterion determined by the Glaucoma VCEP for the *MYOC* gene are in line with those recommended by other VCEPs for similar autosomal dominant heterogeneous diseases such as *MYH7*-associated inherited cardiomyopathies (Kelly et al., 2018), genetic hearing loss (Oza et al., 2018) and myeloid malignancy caused by *RUNX1* variants (Luo et al., 2019). This consistency in approach provides a framework that can be applied to other diseases and genes that have similar characteristics but have not yet had the benefit of dedicated VCEP review and rule specification. This is important, as the workload for defining specific rules for every disease-causing gene is somewhat daunting. The genetic hearing loss VCEP has specified a set of rules to be applied across a range of genetic hearing loss-related genes (Oza et al., 2018), streamlining the process for this group of genes that all have similar characteristics. This approach will be necessary for the efficient specification of rules for other heterogeneous but monogenic diseases.

The Glaucoma VCEP has curated 81 variants using *MYOC-*specific rules within the framework of the ACMG/AMP guidelines and these classifications are available in ClinVar. The next task for the VCEP is to curate the remaining reported variants in *MYOC*. The VCEP will curate novel variants as they are reported. We will also review any variants submitted to ClinVar by one or two star submitters with a different classification to that assigned by the VCEP, as additional evidence may change the classifications. Variants with a medically significant difference (P/LP vs B/LB/VUS) will be reassessed within 3 months of being notified of the discrepant ClinVar classification. Variants classified as LP and VUS will be reviewed every 2 years and LB variants will be reassessed when new large population datasets are released as per ClinGen protocol, to ensure up-to-date information is available for all variants. The VCEP will review its *MYOC-*specified rules every 2 years or sooner as necessary if new knowledge or recommendations from ClinGen arise.

## Data Availability

All data produced in the present study are available upon reasonable request to the authors

## ACKNOWLEDGEMENTS

This work was funded by a National Health and Medical Research Council (NHMRC) of Australia Centre for Research Excellence grant (GNT1116360), Program Grant (GNT1150144), Practitioner Fellowships to J.E.C, D.A.M and A.W.H., The Hospital Research Foundation Early Career Fellowship to E.S, A Core Grant for Vision Research from the National Eye Institute/National Institutes of Health to the University of Wisconsin-Madison (P30EY016665) and an Unrestricted Grant from Research to Prevent Blindness, Inc. to the UW-Madison Department of Ophthalmology and Visual Sciences to T.L.Y. and K.N.W. The authors would like to acknowledge the support of the ClinGen Sequence Variant Interpretation and Ocular Clinical Domain Working Groups, especially Kristy Lee.

