## Supplementary Table 1 and Supplementary Figures for "SPECIFICATIONS OF THE ACMG/AMP VARIANT CURATION GUIDELINES FOR MYOCILIN: RECOMMENDATIONS FROM THE CLINGEN GLAUCOMA EXPERT PANEL"

#### Supplementary Information

### Supplementary Tables

**Supplementary Table 1:** OR (95%CI) and P-value (Fishers Exact Test) for proband counts versus allele counts in gnomad V2.1.1 full population (n=125748) or NFE subset (n=56885). Size of the case cohort is based on ANZRAG (n=3522 in total, or n=3277 for European descent only). ORs of 10, 30 and 100 are represented in yellow, orange and red respectively.

| Number of probands with variant allele | 3522 probands vs 125748 controls 6 alleles present in controls | | | 3277 probands vs 56885 controls 3 alleles present in controls | | |
|---|---|---|---|---|---|---|
| | OR | (95%CI) | P-value | OR | (95%CI) | P-value |
| 2 | 11.91 | (1.2-66.6) | 0.01860 | 11.58 | (1.0-101.2) | 0.02656 |
| 3 | 17.87 | (2.9-83.9) | 0.00150 | 17.37 | (2.3-129.4) | 0.00285 |
| 4 | 23.83 | (4.9-100.8) | 0.00010 | 23.18 | (3.9-158.3) | 0.00027 |
| 5 | 29.79 | (7.2-117.3) | 6.03E-06 | 28.97 | (5.6-186.6) | 2.33E-05 |
| 6 | 35.76 | (9.6-133.8) | 3.27E-07 | 34.77 | (7.4-214.8) | 1.89E-06 |
| 7 | 41.71 | (12.0-150.0) | 1.65E-08 | 40.58 | (9.3-242.8) | 1.47E-07 |
| 8 | 47.69 | (14.5-167.5) | 7.81E-10 | 46.34 | (11.1-270.6) | 1.09E-08 |
| 9 | 53.71 | (17.1-184.1) | 3.53E-11 | 52.18 | (13.0-298.0) | 7.91E-10 |
| 10 | 59.64 | (19.6-200.0) | 1.53E-12 | 58.01 | (14.9-325.1) | 5.58E-11 |
| 11 | 65.75 | (22.2-216.9) | 6.43E-14 | 63.84 | (16.9-353.9) | 3.85E-12 |
| 12 | 71.61 | (24.9-231.3) | 2.62E-15 | 69.54 | (18.8-382.8) | 2.61E-13 |
| 13 | 77.54 | (27.5-249.0) | <2.2E-16 | 75.62 | (20.7-411.9) | 1.74E-14 |
| 14 | 83.48 | (30.2-265.1) | <2.2E-16 | 81.42 | (22.7-441.0) | 1.15E-15 |
| 15 | 89.57 | (32.8-283.8) | <2.2E-16 | 87.23 | (24.6-469.4) | <2.2E-16 |
| 16 | 95.60 | (35.5-296.3) | <2.2E-16 | 93.06 | (26.6-497.7) | <2.2E-16 |
| 17 | 101.61 | (38.2-314.3) | <2.2E-16 | 98.89 | (28.6-525.9) | <2.2E-16 |
| 18 | 107.85 | (40.9-332.1) | <2.2E-16 | 104.72 | (30.5-554.1) | <2.2E-16 |
| 19 | 113.30 | (43.6-349.1) | <2.2E-16 | 110.57 | (32.5-582.4) | <2.2E-16 |
| 20 | 119.41 | (46.4-366.5) | <2.2E-16 | 116.42 | (34.5-610.7) | <2.2E-16 |

**Supplementary Table 2a**: Functional information extracted from the literature for pilot variants.

*See attached table*

**Supplementary Table 2b**: Details of each of the studies included in Supplementary Table 2a.

*See attached table*

**Supplementary Table 3**: Curation of pilot variants indicating variant identification and criteria applied.

*See attached table*

#### Supplementary Figures

**Supplementary Figure 1:** CADD and REVEL scores for missense variants reported in ClinVar as at July 2020. Colours reflect the clinical classification in ClinVar at that time (prior to application of VCEP MYOC specific rules). Variants labelled as “conflicting evidence” are included as VUS in this plot.

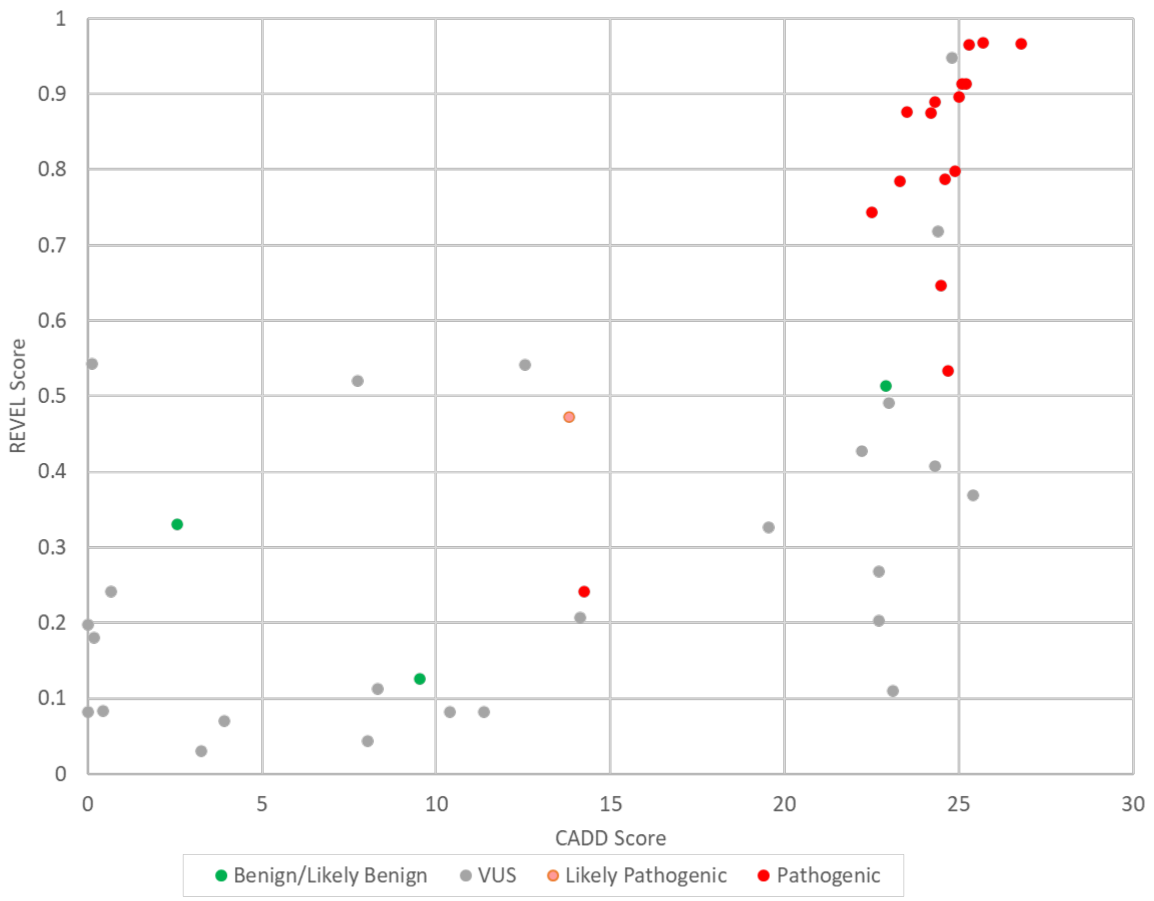

**Supplementary Figure 2:** Tables 1 & 2 from SVI point recommendation for determining the evidence strength level for de novo occurrence (PS2/PM6) (Sequence Variant Interpretation Recommendation for De Novo Criteria (PS2 & PM6) - Version 1.1. ClinGen; 2021.)

**Table 1. Points awarded per de novo occurrence**

| Phenotypic consistency | Points per Proband | |
|---|---|---|
| | Confirmed de novo | Assumed de novo |
| Phenotype highly specific for gene | 2 | 1 |
| Phenotype consistent with gene but not highly specific | 1 | 0.5 |
| Phenotype consistent with gene but not highly specific and high genetic heterogeneity* | 0.5 | 0.25 |
| Phenotype not consistent with gene | 0 | 0 |

*Maximum allowable value of 1 may contribute to overall score

**Table 2. Recommendation for determining the appropriate ACMG/AMP evidence strength level for de novo occurrence(s)**

| **Supporting** (PS2_Supporting or PM6_Supporting) | **Moderate** (PS2_Moderate or PM6) | **Strong** (PS2 or PM6_Strong) | **Very Strong** (PS2_VeryStrong or PM6_VeryStrong) |
|---|---|---|---|
| **0.5** | **1** | **2** | **4** |

**Supplementary Figure 3:** Maximum reported population minor allele frequency in gnomAD plotted against codon number for each pilot variant. The minor allele frequency thresholds employed for each rule are shown and variants are coloured by their final classification.

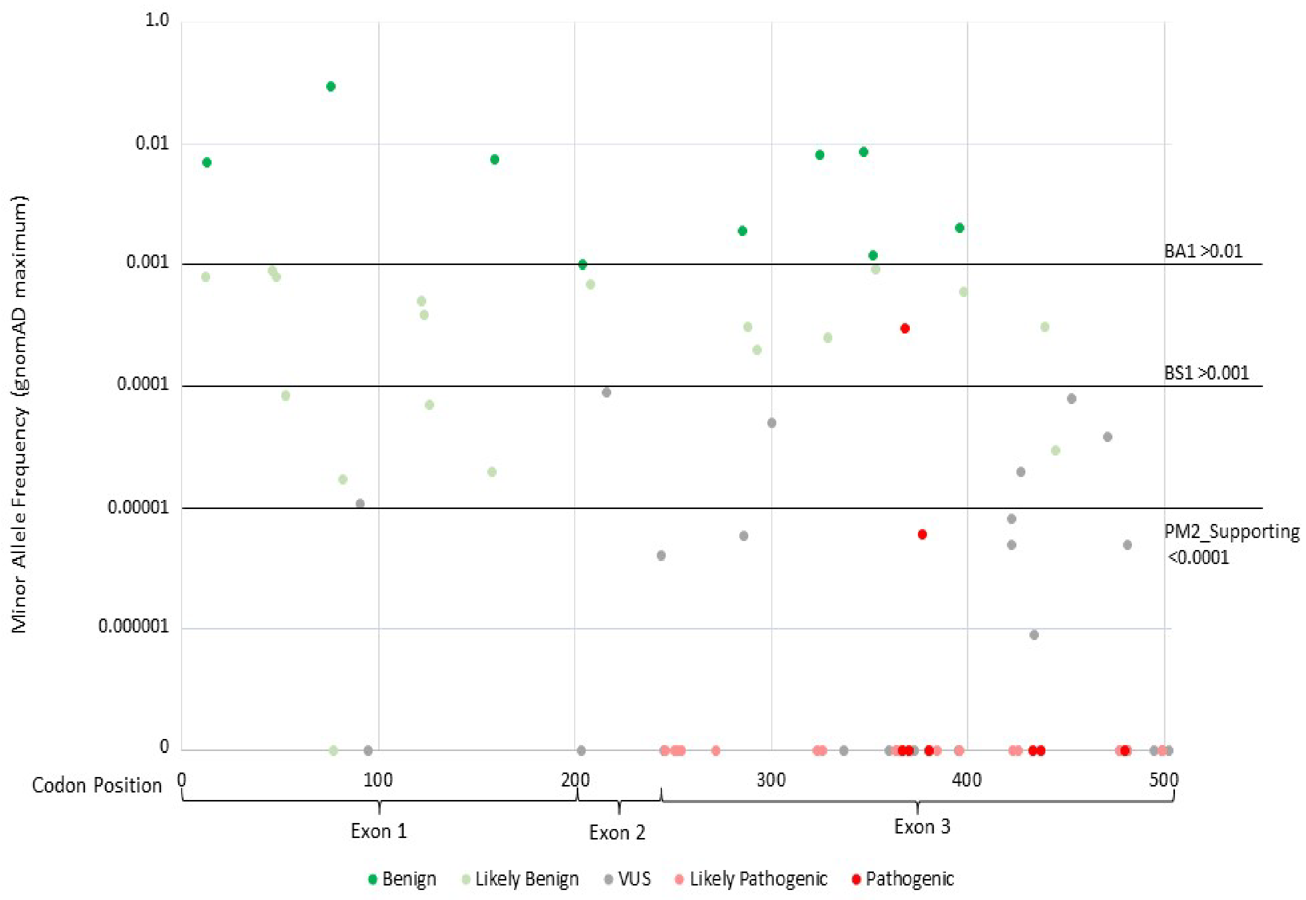
