## Supplementary Table 2a for "SPECIFICATIONS OF THE ACMG/AMP VARIANT CURATION GUIDELINES FOR MYOCILIN: RECOMMENDATIONS FROM THE CLINGEN GLAUCOMA EXPERT PANEL"

Supplementary Table 2a: Functional information extracted from the literature for pilot variants. Type of assay and first author are listed at the top, along with the level of BS3 or PS3 able to be applied based on OddsPath ratio. Assay results for each variant, the level applied and the final classification considering all rules are given. See Supplementary Table 2b for details of each study. na=not applied due to conflicting results or not meeting required OddsPath ratio. *data received from John Hulleman (personal communication) using same methods as Nakahara

| | Nakahara | | | Zadoo | Gobeil | Zhou | Shimizu | Liu | Vollrath | | Jacobson | Izumi | Jia | | |
|---|---|---|---|---|---|---|---|---|---|---|---|---|---|---|---|
| | Secretion eGLuc2 | Secretion (FLAG) | Solubility (FLAG) | Secretion eGLuc2 | Secretion | Solubility | Solubility | Solubility | Solubility | Secretion | Secretion | Secretion | Solubility | | |
| **PS3** | PS3_Moderate | | | not applied | PS3_Moderate | PS3_Supporting | | not applied | not applied | | not applied | not applied | not applied | | |
| **BS3** | BS3_Moderate | | | not applied | BS3_Moderate | BS3_Moderate | | not applied | not applied | | BS3_Supporting | not applied | not applied | | |
| **Variant** | | | | | | | | | | | | | | **PS3/BS3 applied** | **Final Classification** |
| G12R | secreted | secreted | soluble | | | | | | | | | | | BS3_Moderate | LB |
| Q48H | secreted | secreted | soluble | | secreted | | | | | | | | | BS3_Moderate | LB |
| V53A | secreted | secreted | soluble | | | | | | | | | | | BS3_Moderate | LB |
| R76K | secreted | secreted | soluble | | secreted | | | | | | | | | BS3_Moderate | B |
| R82C | | | | | secreted | | | | | | | | | BS3_Moderate | LB |
| L95P | | | | | secreted | | | | | | | | | BS3_Moderate | VUS |
| R126W | | | | | secreted | | | | | | | | | BS3_Moderate | LB |
| R158Q | | | | | secreted | | | | | | | secreted | | BS3_Moderate | LB |
| S203F | | | | | secreted | | | | | | | | | BS3_Moderate | VUS |
| T204T | secreted | secreted | soluble | | | | | | | | | | | BS3_Moderate | B |
| D208E | | | | secreted | | | | | | | | secreted | | na | LB |
| G244V | | | | secreted | | | | | | | | | | na | VUS |
| C245Y | | | | non secreted | | | | | | | | | insoluble | na | VUS |
| G246R | | | | non secreted | non secreted | | | | insoluble | non secreted | | | | PS3_Moderate | LP |
| G252R | | | | | non secreted | | insoluble | | | | | | | PS3_Moderate | LP |
| P254L | non secreted | non secreted | insoluble | | | | | | | | | | | PS3_Moderate | LP |
| R272G | | | | | | | insoluble | | insoluble | non secreted | | | | PS3_Supporting | VUS |
| W286R | | | | | non secreted | | | | | | | | | PS3_Moderate | VUS |
| T293K | | | | | secreted | | | | | | | | | BS3_Moderate | LB |
| E300K | | | | non secreted | | | | | | | | | | na | VUS |
| E323K | | | | | non secreted | insoluble | insoluble | insoluble | insoluble | partial | | | | PS3_Moderate | LP |
| T325T | secreted | secreted | soluble | | | | | | | | | | | BS3_Moderate | B |
| G326R | | | | | non secreted | | | | | | | | | PS3_Moderate | LP |
| V329M | | | | | | | soluble | | | | | | | BS3_Moderate | LB |
| E352K | | | | | secreted | soluble | | | | | | | | BS3_Moderate | B |
| T353I | | | | | secreted | | | | | | | | | BS3_Moderate | LB |
| I360N | | | | | | | | | | | | non secreted | | na | VUS |
| A363T* | | non secreted | insoluble | | | | | | | | | non secreted | | PS3_Moderate | LP |
| G364V | | | | | non secreted | partial | | insoluble | partial | partial | partial | | | PS3_Moderate | LP |
| G367R | non secreted | non secreted | insoluble | | non secreted | | | | | | | | | PS3_Moderate | P |
| G367_Q368delinsV | | | | | | insoluble | | | insoluble | non secreted | | | | PS3_Supporting | LP |
| Q368X | | | | | non secreted | insoluble | insoluble | | | | non secreted | | | PS3_Moderate | P |
| P370L | | | | | non secreted | insoluble | insoluble | insoluble | insoluble | non secreted | | | insoluble | PS3_Moderate | P |
| T377M | | | | | partial | partial | insoluble | | partial | partial | | | | na | P |
| D380H | secreted | non secreted | insoluble | | | | | | | | | | | PS3_Moderate | LP |
| D380Y* | | non secreted | insoluble | | | | | | | | | | | PS3_Moderate | LP |
| D380A | | | | | non secreted | partial | | insoluble | partial | partial | | | | PS3_Moderate | P |
| D380G* | | non secreted | insoluble | | | | | | | | | | | PS3_Moderate | VUS |
| D384N | | | | | | | | | | | | | insoluble | na | LP |
| D395_E396insDP | non secreted | non secreted | insoluble | | | | | | | | | | | PS3_Moderate | VUS |
| E396dup | non secreted | non secreted | insoluble | | non secreted | | | | | | | | | PS3_Moderate | LP |
| K398R | | | | | secreted | soluble | soluble | | | | secreted | | | BS3_Moderate | LB |
| R422C | | | | | | partial | | | | | | | | na | VUS |

| | Nakahara | | | Zadoo | Gobeil | Zhou | Shimizu | Liu | Vollrath | | Jacobson | Izumi | Jia | | |
|---|---|---|---|---|---|---|---|---|---|---|---|---|---|---|---|
| | Secretion eGLuc2 | Secretion (FLAG) | Solubility (FLAG) | Secretion eGLuc2 | Secretion | Solubility | Solubility | Solubility | Solubility | Secretion | Secretion | Secretion | Solubility | | |
| **PS3** | PS3_Moderate | | | not applied | PS3_Moderate | PS3_Supporting | | not applied | not applied | | not applied | not applied | not applied | | |
| **BS3** | BS3_Moderate | | | not applied | BS3_Moderate | BS3_Moderate | | not applied | not applied | | BS3_Supporting | not applied | not applied | | |
| **Variant** | | | | | | | | | | | | | | **PS3/BS3 applied** | **Final Classification** |
| R422H | | | | | | soluble | | | | | | | | BS3_Moderate | VUS |
| K423E | | | | | non secreted | insoluble | | insoluble | insoluble | non secreted | non secreted | | | PS3_Moderate | LP |
| V426F | | | | | non secreted | insoluble | insoluble | | insoluble | partial | | | | PS3_Moderate | LP |
| A427T | | | | | secreted | | | | | | | | | BS3_Moderate | VUS |
| C433R | | | | | non secreted | | | | | | | | | PS3_Moderate | P |
| G434S | | | | secreted | | | | | | | | | | na | VUS |
| Y437H | non secreted | non secreted | insoluble | non secreted | non secreted | insoluble | | insoluble | insoluble | non secreted | non secreted | | insoluble | PS3_Moderate | P |
| A445V | secreted | secreted | soluble | | secreted | | | | | | | | | BS3_Moderate | LB |
| Y471C | | | | secreted | | | | | | | | | | na | VUS |
| I477N | | | | non secreted | non secreted | insoluble | insoluble | | insoluble | non secreted | | | | PS3_Moderate | LP |
| I477S | | | | | | insoluble | | | insoluble | non secreted | non secreted | non secreted | | PS3_Supporting | VUS |
| N480K (c.1440C>A) | non secreted | non secreted | insoluble | | non secreted | insoluble | | | insoluble | partial | | | | PS3_Moderate | P |
| N480K (c.1440C>G) | | | | | | | | | | | | | | | LP |
| P481S | secreted | secreted | insoluble | | | | | | | | | | | na | VUS |
| P481L | | | | | non secreted | | | | | | | | | PS3_Moderate | LP |
| V495I | | | | | | soluble | | | | | | | | BS3_Moderate | VUS |
| I499F | | | | | non secreted | partial | | | partial | partial | | | | PS3_Moderate | LP |
| I499S | | | | | | | insoluble | | | | | | | PS3_Supporting | VUS |
| S502P | | | | | non secreted | | | | | | | | | PS3_Moderate | VUS |
