## Supplementary Table 2b for "SPECIFICATIONS OF THE ACMG/AMP VARIANT CURATION GUIDELINES FOR MYOCILIN: RECOMMENDATIONS FROM THE CLINGEN GLAUCOMA EXPERT PANEL"

Supplementary Table 2b: Details of each of the studies included in Supplementary Table 2a

| PMID | BioArxiv preprint | 27092720 | 16466712 | 10545602 | 11004290 | 15069026 | 16297911 | | 11152659 | 14688426 | 19234343 |
|---|---|---|---|---|---|---|---|---|---|---|---|
| DOI / link | https://www.biorxiv.org/content/10.1101/2021.10.30.466573v1 | https://doi.org/10.1167/iovs.15-18789 | https://doi.org/10.1016/j.exer.2005.11.002 | https://doi.org/10.1093/hmg/8.12.2221 | https://doi.org/10.1016/s0002-9394(00)00536-5 | https://doi.org/10.1093/hmg/ddh128 | https://doi.org/10.1016/j.exer.2005.10.007 | | https://doi.org/10.1093/hmg/10.2.117 | https://doi.org/10.1159/000074075 | https://doi.org/10.1167/iovs.08-3151 |
| Author | Nakahara | Zadoo | Gobeil | Zhou | Shimizu | Liu | Vollrath | | Jacobson | Izumi | Jia |
| Year | 2021 | 2016 | 2006 | 1999 | 2000 | 2004 | 2006 | | 2001 | 2003 | 2009 |
| **Assay (general description)** | secretion/solubility assays | Secretion assay | Secretion assay | solubility assay | | secretion/solubility assays | | | Secretion assay | Secretion assay | Secretion assay |
| **Cell lines used** | HEK-293A | HEK-293T | COS-7 , iHTM | HEK, COS7, NIH 3T3 and HeLa (most in HEK) | HEK-293T | HEK-293, HTM | HEK-293 | HEK-293 | A549, Cos-7, TM5 | COS-1 | HTM |
| **Promoter** | CMV | CMV | CMV | CMV | unknown, assume CMV | CMV | CMV | CMV | RSV (adenovirus), CMV (transient transfection) | CMV | CMV |
| **Method of overexpression** | transient transfection | transient transfection | transient transfection | transient transfection | transient transfection | transient transfection, adenovirus transduction | transient transfection | transient transfection | adenovirus transduction, transient transfection | transient electroporation | transient transfection, possibly stable cells |
| **Epitope tag** | FLAG | Gaussia luciferase (Gluc), FLAG | untagged, myc | untagged, FLAG | FLAG | FLAG | FLAG | | untagged, FLAG | untagged | 3xFLAG myc |
| **Intracellular protein extraction** | Triton X-100 insoluble (0.1%) | intracellular analysis of luciferase-tagged constructs was not performed | Triton X-100 soluble (0.5%) | Triton X-100 insoluble (0.5%) | Triton X-100 insoluble (0.5%) | Triton-X soluble and insoluble proteins (1%) | Triton-X soluble and insoluble proteins (1%) | | Not specifically mentioned, likely used M-PER, no enrichment of insoluble portion | M-PER soluble | Triton X-100-soluble and insoluble fractions (0.5%) |
| **Detection method** | immunoblot with a rabbit anti-FLAG polyclonal antibody (Invitrogen) | GLuc assay | immunoblot for untagged or myc-tagged myocilin cDNAs, Western blot using custom rabbit anti-MYOC antibody | immunoblot with a M2 anti-FLAG monoclonal antibody (Eastman Kodak) or custom made anti-MYOC polyclonal antibody | FLAG epitope-tagged versions of normal or mutant, immunoblot with an anti-FLAG monoclonal antibody | immunoblot analysis with a monoclonal anti-FLAG antibody | immunoblot for FLAG-epitope-tagged wildtype or various mutant myocilins | immunoblot | immunoblot for untagged and FLAG-tagged MYOC, Western immunoblot using a custom rabbit anti-MYOC peptide antibody 129 (untagged) or mouse M2 anti-FLAG (Sigma) | immunoblot of untagged MYOC using anti-human MYOC peptide antibody 4381R | immunoblot of FLAG epitope using anti-FLAG HRP (Sigma) |
| **Biological replicates (met/not met)** | performed the experiments up to 6 times | performed the experiments up to 8 times | unknown | comparable results have been obtained in more than 20 replications of some experiments | unknown | unknown | transfection and immunoblot process was repeated 2–3 times for each mutant to assess reproducibility | | unknown | unknown | unknown |
| **Technical replicates (met/not met); description** | not met: used singlet data points for each bioligical replicate | not met: used singlet data points for each bioligical replicate | unknown | unknown | unknown | unknown | unknown | | unknown | unknown | unknown |
| **Basic positive control (met/not met); description** | met: wild-type | met: wild-type | met: wild-type | met: wild-type and human to murine substitution (Ala386Ser) | met: wild-type | met: wild-type and human to murine substitution (Ala386Ser) | met: wild-type | | met: wild-type | met: wild type | met: wild-type |
| **Basic negative control (met/not met); description** | met: eGFP | met: untransfected cells | met: empty plasmid | ?met | ?met | ?met | ?met | | met: *lacZ* gene | ?not met | met: empty vector |
| **Validation controls P/LP (#)** | 3 | 2 | 13 | 11 | 6 | 5 | 10 | | 4 | 0 | 3 |
| **Validation controls B/LB (#)** | 5 | 1 | 6 | 2 | 2 | 0 | 0 | | 1 | 1 | 0 |
| **Statistical analysis (general description)** | ImageQuant quantitation values, 1-sample t-test or 2-sample t-test, where appropriate. Significance was set at *P < 0.05, **P < 0.01, and ***P < 0.001 | luminescence values, 1-sample t-test or 2-sample t-test, where appropriate. Significance was set at *P < 0.05, **P < 0.01, and ***P < 0.001 | not performed | not performed | not performed | not performed | not performed | | not performed | not performed | Quantity One image analysis software used, but no statistical test performed |
| **Threshold for normal/abnormal readout** | statistical signifcance vs. controls | statistical signifcance vs. controls | qualitative observation | qualitative observation | qualitative observation | qualitative observation | semi-quantitative based on several immunoblots | | qualitative observation | qualitative observation | qualitative observation |
| **Approved assay (y/n)** | Y | Y | Y | Y | | Y | | | Y | Y | Y |
| **Proposed strength** | Apply PS3_Moderate (OddsPath=8, meets the >4.3 threshold), apply BS3_Moderate (OddsPath=0.077, meets the <0.23 threshold) | Do not apply PS3 (OddsPath=1, does not meet the >2.1 threshold for PS3_Sup), or BS3 (OddsPath=0.5, does not meet the <0.48 threshold for BS3_Sup) | Apply PS3_Moderate (OddsPath=6, meets the >4.3 threshold), apply BS3_Moderate (OddsPath=0.077, meets the <0.23 threshold) | Apply PS3_Supporting (OddsPath=3.0, meets the >2.1 threshold), apply BS3_Moderate (OddsPath=0.083, meets the <0.23 threshold) | | Do not apply PS3 (OddsPath=0, does not meet the >2.1 threshold for PS3_Sup), or BS3 (OddsPath n/a, does not meet the <0.48 threshold for BS3_Sup) | | | Do not apply PS3 (OddsPath = 1, does not meet the >2.1 threshold for PS3_Sup), apply BS3_Supporting (OddsPath=0.25, meets the <0.48 threshold) | Do not apply PS3 (OddsPath n/a, does not meet the >2.1 threshold for PS3_Sup), or BS3 (OddsPath=1, does not meet the <0.48 threshold for BS3_Sup) | Do not apply PS3 (OddsPath=0, does not meet the >2.1 threshold for PS3_Sup), or BS3 (OddsPath n/a, does not meet the <0.48 threshold for BS3_Sup) |
