## Supplementary Table 3 for "SPECIFICATIONS OF THE ACMG/AMP VARIANT CURATION GUIDELINES FOR MYOCILIN: RECOMMENDATIONS FROM THE CLINGEN GLAUCOMA EXPERT PANEL"

Supplementary Table 3: Curation of pilot variants indicating variant identification and criteria applied.

| Genomic (NC_000001.10) | cDNA (NM_000261.2) | Amino acid (NP_000252.1) | Exon | Type | ClinVar ID | CA ID | PS1 | PS2 | PS3 | PS4 | PP1_Str | PM4 | PM5 | PS1_Mod | PS2_Mod | PS3_Mod | PS4_Mod | PP1_Mod | PP1 | PP3 | PS2_Sup | PS3_Sup | PS4_Sup | PM2_Sup | PM4_Sup | PM5_Sup | BA1 | BS1 | BS3_Mod | BP4 | BP7 | BS3_Sup | Summary of criteria applied | Score | Final classification | ClinVar classification |
|---|---|---|---|---|---|---|---|---|---|---|---|---|---|---|---|---|---|---|---|---|---|---|---|---|---|---|---|---|---|---|---|---|---|---|---|---|
| 171621718C>G | 34G>C | Gly12Arg | 1 | Missense | 876089 | CA1244345 | | | | | | | | | | | | | | | | | | | | | | ✓ | ✓ | | | | BS1, BS3_Mod | -6 | LB | B/LB |
| 171621713A>C | 39T>G | Pro13Pro | 1 | Synonymous | 293724 | CA1244342 | | | | | | | | | | | | | | | | | | | | | ✓ | | | ✓ | ✓ | | BA1, BP4, BP7 | | B | B |
| 171621616G>A | 136C>T | Arg46Ter | 1 | Nonsense | 7955 | CA119179 | | | | | | | | | | | | | | | | | | | | | | ✓ | | | | | BS1 | -4 | LB | B/LB |
| 171621608C>A | 144G>T | Gln48His | 1 | Missense | 7958 | CA119183 | | | | | | | | | | | | | | | | | | | | | | ✓ | ✓ | | | | BS1, BS3_Mod | -6 | LB | B/P (conflicting) |
| 171621594A>G | 158T>C | Val53Ala | 1 | Missense | 631580 | CA1244317 | | | | | | | | | | | | | | | | | | | | | | | ✓ | | | | BS3_Mod | -2 | LB | VUS |
| 171621525C>T | 227G>A | Arg76Lys | 1 | Missense | 193066 | CA200294 | | | | | | | | | | | | | | | | | | | | | ✓ | | ✓ | | | | BA1, BS3_Mod | | B | B/LB/VUS (conflicting) |
| 171621508G>A | 244C>T | Arg82Cys | 1 | Missense | 806285 | CA1244309 | | | | | | | | | | | | | | | | | | | | | | | ✓ | | | | BS3_Mod | -2 | LB | VUS |
| 171621481G>A | 271C>T | Arg91Ter | 1 | Nonsense | | CA1244303 | | | | | | | | | | | | | | | | | | | | | | | | | | | | 0 | VUS | |
| 171621468A>G | 284T>C | Leu95Pro | 1 | Missense | | CA343718858 | | | | | | | | | | | | | | | | | | ✓ | | | | | ✓ | | | | PM2_Sup, BS3_Mod | -1 | VUS | |
| 171621386G>A | 366C>T | Gly122Gly | 1 | Synonymous | 193067 | CA238570 | | | | | | | | | | | | | | | | | | | | | | ✓ | | ✓ | ✓ | | BS1, BP4, BP7 | -6 | LB | B/VUS (conflicting) |
| 171621383G>A | 369C>T | Thr123Thr | 1 | Synonymous | 874205 | CA1244290 | | | | | | | | | | | | | | | | | | | | | | ✓ | | ✓ | | | BS1, BP4 | -5 | LB | VUS |
| 171621376G>A | 376C>T | Arg126Trp | 1 | Missense | 1303146 | CA1244289 | | | | | | | | | | | | | | | | | | | | | | | ✓ | | | | BS3_Mod | -2 | LB | VUS |
| 171621279C>T | 473G>A | Arg158Gln | 1 | Missense | 874204 | CA1244272 | | | | | | | | | | | | | | | | | | | | | | | ✓ | | | | BS3_Mod | -2 | LB | VUS |
| 171621275T>C | 477A>G | Leu159Leu | 1 | Synonymous | 293717 | CA1244271 | | | | | | | | | | | | | | | | | | | | | ✓ | | | ✓ | | | BA1, BP4 | | B | B |
| 171607859G>A | 608C>T | Ser203Phe | 2 | Missense | | CA343727142 | | | | | | | | | | | | | | | | | | ✓ | | | | | ✓ | | | | PM2_Sup, BS3_Mod | -1 | VUS | |
| 171607855C>A | 612G>T | Thr204Thr | 2 | Synonymous | 728934 | CA1244212 | | | | | | | | | | | | | | | | | | | | | ✓ | | ✓ | ✓ | ✓ | | BA1, BS3_Mod, BP4, BP7 | | B | B/LB |
| 171607843G>C | 624C>G | Asp208Glu | 2 | Missense | 876020 | CA1244210 | | | | | | | | | | | | | | | | | | | | | | ✓ | | ✓ | | | BS1, BP4 | -5 | LB | B |
| 171607819C>T | 648G>A | Lys216Lys | 2 | Synonymous | 293715 | CA1244204 | | | | | | | | | | | | | | | | | | | | | | | | | | | | 0 | VUS | B/VUS (conflicting) |
| 171605849C>A | 731G>T | Gly244Val | 3 | Missense | | CA1244164 | | | | | | | | | | | | | | ✓ | | | ✓ | ✓ | | | | | | | | | PP3, PS4_Sup, PM2_Sup | 3 | VUS | |
| 171605846C>T | 734G>A | Cys245Tyr | 3 | Missense | 7959 | CA119184 | | | | | | | | | | | | | ✓ | ✓ | | | | ✓ | | | | | | | | | PP1, PP3, PM2_Sup | 3 | VUS | P |
| 171605844C>T | 736G>A | Gly246Arg | 3 | Missense | | CA343726300 | | | | | | | | | | ✓ | | ✓ | | ✓ | | | | ✓ | | | | | | | | | PS3_Mod, PP1_Mod, PP3, PM2_Sup | 6 | LP | |
| 171605828A>G | 752T>C | Val251Ala | 3 | Missense | | CA343726262 | | | | | ✓ | | | | ✓ | | | | | ✓ | | | ✓ | ✓ | | | | | | | | | PP1_Str, PS2_Mod, PP3, PS4_Sup, PM2_Sup | 9 | LP | |
| 171605826C>T | 754G>A | Gly252Arg | 3 | Missense | 7960 | CA119185 | | | | | ✓ | | | | | ✓ | | | | ✓ | | | ✓ | ✓ | | | | | | | | | PP1_Str, PS3_Mod, PP3, PS4_Sup, PM2_Sup | 9 | LP | P |
| 171605819G>A | 761C>T | Pro254Leu | 3 | Missense | | CA343726245 | | | | | | | | | ✓ | ✓ | | | | ✓ | | | | ✓ | | | | | | | | | PS2_Mod, PS3_Mod, PP3, PM2_Sup | 6 | LP | |
| 171605766G>C | 814C>G | Arg272Gly | 3 | Missense | | CA343726016 | | | | | | | | | | | | ✓ | | ✓ | | ✓ | | ✓ | | | | | | | | | PP1_Mod, PP3, PS3_Sup, PM2_Sup | 5 | VUS | |
| 171605725C>A | 855G>T | Thr285Thr | 3 | Synonymous | 196241 | CA202234 | | | | | | | | | | | | | | | | | | | | | ✓ | | | ✓ | ✓ | | BA1, BP4, BP7 | | B | B/LB/VUS (conflicting) |
| 171605724A>G | 856T>C | Trp286Arg | 3 | Missense | | CA343725818 | | | | | | | | | | ✓ | | | | ✓ | | | ✓ | ✓ | | | | | | | | | PS3_Mod, PP3, PS4_Sup, PM2_Sup | 5 | VUS | |
| 171605716G>A | 864C>T | Ile288Ile | 3 | Synonymous | 874154 | CA1244136 | | | | | | | | | | | | | | | | | | | | | | ✓ | | ✓ | ✓ | | BS1, BP4, BP7 | -6 | LB | B/LB |
| 171605702G>T | 878C>A | Thr293Lys | 3 | Missense | 876939 | CA1244131 | | | | | | | | | | | | | | | | | | | | | | ✓ | ✓ | | | | BS1, BS3_Mod | -6 | LB | VUS |
| 171605682C>T | 898G>A | Glu300Lys | 3 | Missense | | CA1244125 | | | | | | | | | | | | | | ✓ | | | | | | | | | | | | | PP3 | 1 | VUS | |
| 171605613C>T | 967G>A | Glu323Lys | 3 | Missense | | CA343725312 | | | | | | | | | | ✓ | | ✓ | | ✓ | | | ✓ | ✓ | | | | | | | | | PS3_Mod, PP1_Mod, PP3, PS4_Sup, PM2_Sup | 7 | LP | |
| 171605605C>T | 975G>A | Thr325Thr | 3 | Synonymous | 293711 | CA1244108 | | | | | | | | | | | | | | | | | | | | | ✓ | | ✓ | ✓ | ✓ | | BA1, BS3_Mod, BP4, BP7 | | B | B |
| 171605604C>G | 976G>C | Gly326Arg | 3 | Missense | | CA343725268 | | | | | | | | | | ✓ | | ✓ | | ✓ | | | | ✓ | | | | | | | | | PS3_Mod, PP1_Mod, PP3, PM2_Sup | 6 | LP | |
| 171605595C>T | 985G>A | Val329Met | 3 | Missense | 111192 | CA1244106 | | | | | | | | | | | | | | | | | | | | | | ✓ | ✓ | | | | BS1, BS3_Mod | -6 | LB | LB |
| 171605571delC | 1009delC | Gln337ArgfsTer9 | 3 | Frameshift | | CA1139655085 | | | | | | ✓ | | | | | | ✓ | | | | | | ✓ | | | | | | | | | PM4, PP1_Mod, PM2_Sup | 5 | VUS | |
| 171605539A>G | 1041T>C | Tyr347Tyr | 3 | Synonymous | 293709 | CA1244091 | | | | | | | | | | | | | | | | | | | | | ✓ | | | ✓ | ✓ | | BA1, BP4, BP7 | | B | B/LB |
| 171605526C>T | 1054G>A | Glu352Lys | 3 | Missense | 875955 | CA1244087 | | | | | | | | | | | | | | | | | | | | | ✓ | | ✓ | | | | BA1, BS3_Mod | | B | B |
| 171605522G>A | 1058C>T | Thr353Ile | 3 | Missense | 1013538 | CA1244086 | | | | | | | | | | | | | | | | | | | | | | ✓ | ✓ | | | | BS1, BS3_Mod | -6 | LB | VUS |
| 171605501A>T | 1079T>A | Ile360Asn | 3 | Missense | | CA343724796 | | | | | | | | | | | | | | | | | | ✓ | | | | | | | | | PM2_Sup | 1 | VUS | |
| 171605493C>T | 1087G>A | Ala363Thr | 3 | Missense | | CA343724770 | | | | | | | | | | ✓ | | ✓ | | ✓ | | | ✓ | ✓ | | | | | | | | | PS3_Mod, PP1_Mod, PP3, PS4_Sup, PM2_Sup | 7 | LP | |
| 171605489C>A | 1091G>T | Gly364Val | 3 | Missense | 7947 | CA119170 | | | | | ✓ | | | | | ✓ | | | | ✓ | | | ✓ | ✓ | | | | | | | | | PP1_Str, PS3_Mod, PP3, PS4_Sup, PM2_Sup | 9 | LP | P |
| 171605481C>T | 1099G>A | Gly367Arg | 3 | Missense | 7952 | CA119176 | | | | ✓ | ✓ | | | | | ✓ | | | | ✓ | | | | ✓ | | | | | | | | | PS4, PP1_Str, PS3_Mod, PP3, PM2_Sup | 12 | P | P |
| 171605477_171605480delinsA | 1100_1103delinsT | Gly367_Gln368delinsVal | 3 | Inframe del/ins | | CA1139532761 | | | | | ✓ | | | | | | | | | | | ✓ | ✓ | ✓ | ✓ | | | | | | | | PP1_Str, PS3_Sup, PS4_Sup, PM2_Sup, PM4_Sup | 8 | LP | |
| 171605478G>A | 1102C>T | Gln368Ter | 3 | Nonsense | 7949 | CA119172 | | | | ✓ | ✓ | ✓ | | | | ✓ | | | | | | | | | | | | | | | | | PS4, PP1_Str, PM4, PS3_Mod | 12 | P | P/LP |
| 171605471G>A | 1109C>T | Pro370Leu | 3 | Missense | 7948 | CA119171 | | | | ✓ | ✓ | | | | | ✓ | | | | ✓ | | | | ✓ | | | | | | | | | PS4, PP1_Str, PS3_Mod, PP3, PM2_Sup | 12 | P | P |
| 171605461C>T | 1119G>A | Trp373Ter | 3 | Nonsense | | CA32685688 | | | | | | ✓ | | | | | | | | | | | | ✓ | | | | | | | | | PM4, PM2_Sup | 3 | VUS | |
| 171605450G>A | 1130C>T | Thr377Met | 3 | Missense | | CA1244075 | | | | ✓ | ✓ | | | | | | | | | ✓ | | | | ✓ | | | | | | | | | PS4, PP1_Str, PP3, PM2_Sup | 10 | P | |
| 171605442C>G | 1138G>C | Asp380His | 3 | Missense | 7961 | CA119186 | | | | | | | | | | ✓ | | ✓ | | ✓ | | | | ✓ | | | | | | | | | PS3_Mod, PP1_Mod, PP3, PM2_Sup | 6 | LP | P |
| 171605442C>T | 1138G>A | Asp380Asn | 3 | Missense | | CA343724574 | | | | | | | | | | | | | | ✓ | | | ✓ | ✓ | | | | | | | | | PP3, PS4_Sup, PM2_Sup | 3 | VUS | |
| 171605442C>A | 1138G>T | Asp380Tyr | 3 | Missense | | CA343724573 | | | | | | | ✓ | | | ✓ | | | | ✓ | | | | ✓ | | | | | | | | | PM5, PS3_Mod, PP3, PM2_Sup | 6 | LP | |
| 171605441T>G | 1139A>C | Asp380Ala | 3 | Missense | | CA343724571 | | | | | ✓ | | | | | ✓ | ✓ | | | ✓ | | | | ✓ | | ✓ | | | | | | | PP1_Str, PS3_Mod, PS4_Mod, PP3, PM2_Sup, PM5_Sup | 11 | P | |
| 171605441T>C | 1139A>G | Asp380Gly | 3 | Missense | | CA343724569 | | | | | | | | | | ✓ | | | | ✓ | | | | ✓ | | ✓ | | | | | | | PS3_Mod, PP3, PM2_Sup, PM5_Sup | 5 | VUS | |
| 171605430C>T | 1150G>A | Asp384Asn | 3 | Missense | | CA343724529 | | | | | ✓ | | | | | | | | | ✓ | | | ✓ | ✓ | | | | | | | | | PP1_Str, PS4_Sup, PP3, PM2_Sup | 7 | LP | |
| 171605392_171605393insTCTGG | 1187_1188insCCCAGA | Asp395_Glu396insAspPro | 3 | Inframe del/ins | | CA1139654950 | | | | | | | | | | ✓ | | | ✓ | | | | | ✓ | ✓ | | | | | | | | PS3_Mod, PP1, PM2_Sup, PM4_Sup | 5 | VUS | |
| 171605392_171605394dup | 1187_1189dup | Glu396dup | 3 | Inframe del/ins | | CA1139532728 | | | | | | | | | | ✓ | | ✓ | | | | | | ✓ | ✓ | | | | | | | | PS3_Mod, PP1_Mod, PM2_Sup, PM4_Sup | 6 | LP | |
| 171605392C>T | 1188G>A | Glu396Glu | 3 | Synonymous | 293708 | CA1244067 | | | | | | | | | | | | | | | | | | | | | ✓ | | | ✓ | ✓ | | BA1, BP4, BP7 | | B | B/LB |
| 171605387T>C | 1193A>G | Lys398Arg | 3 | Missense | 403218 | CA1244064 | | | | | | | | | | | | | | | | | | | | | | ✓ | ✓ | | | | BS1, BS3_Mod | -6 | LB | B/LB/VUS (conflicting) |
| 171605316G>A | 1264C>T | Arg422Cys | 3 | Missense | | CA1244053 | | | | | | | | | | | | | | | | | | ✓ | | | | | | | | | PM2_Sup | 1 | VUS | |
| 171605315C>T | 1265G>A | Arg422His | 3 | Missense | | CA1244052 | | | | | | | | | | | | | | ✓ | | | | ✓ | | | | | ✓ | | | | PP3, PM2_Sup, BS3_Mod | 0 | VUS | |
| 171605313T>C | 1267A>G | Lys423Glu | 3 | Missense | 7954 | CA119178 | | | | | ✓ | | | | | ✓ | | | | ✓ | | | ✓ | ✓ | | | | | | | | | PP1_Str, PS3_Mod, PP3, PS4_Sup, PM2_Sup | 9 | LP | P |
| 171605304C>A | 1276G>T | Val426Phe | 3 | Missense | | CA343724072 | | | | | ✓ | | | | | ✓ | | | | ✓ | | | ✓ | ✓ | | | | | | | | | PP1_Str, PS3_Mod, PP3, PS4_Sup, PM2_Sup | 9 | LP | |
| 171605301C>T | 1279G>A | Ala427Thr | 3 | Missense | | CA1244050 | | | | | | | | | | | | | | ✓ | | | | | | | | | ✓ | | | | PP3, BS3_Mod | -1 | VUS | |
| 171605283A>G | 1297T>C | Cys433Arg | 3 | Missense | 7956 | CA119181 | | | | ✓ | ✓ | | | | | ✓ | | | | ✓ | | | | ✓ | | | | | | | | | PS4, PP1_Str, PS3_Mod, PP3, PM2_Sup | 12 | P | P |
| 171605280C>T | 1300G>A | Gly434Ser | 3 | Missense | | CA343723925 | | | | | | | | | | | | | | ✓ | | | | ✓ | | | | | | | | | PP3, PM2_Sup | 2 | VUS | |
| 171605271A>G | 1309T>C | Tyr437His | 3 | Missense | 7946 | CA119169 | | | | | ✓ | | | | | ✓ | ✓ | | | ✓ | | | | ✓ | | | | | | | | | PP1_Str, PS3_Mod, PS4_Mod, PP3, PM2_Sup | 10 | P | P |
| 171605263G>A | 1317C>T | Val439Val | 3 | Synonymous | 806281 | CA1244042 | | | | | | | | | | | | | | | | | | | | | | ✓ | | ✓ | | | BS1, BP4 | -5 | LB | LB |
| 171605246G>A | 1334C>T | Ala445Val | 3 | Missense | 631578 | CA1244040 | | | | | | | | | | | | | | | | | | | | | | | ✓ | | | | BS3_Mod | -2 | LB | LP |
| 171605223del | 1357delT | Tyr453MetfsTer11 | 3 | Frameshift | | CA1244034 | | | | | | | | | | | | | | | | | | | ✓ | | | | | | | | PM4_Sup | 1 | VUS | |
| 171605168T>C | 1412A>G | Tyr471Cys | 3 | Missense | | CA1244027 | | | | | | | | | | | | | | ✓ | | | | | | | | | | | | | PP3 | 1 | VUS | |
| 171605150A>T | 1430T>A | Ile477Asn | 3 | Missense | 30205 | CA129023 | | | | | ✓ | | | | | ✓ | | | | | | | ✓ | ✓ | | | | | | | | | PP1_Str, PS3_Mod, PS4_Sup, PM2_Sup | 8 | LP | P |
| 171605150A>C | 1430T>G | Ile477Ser | 3 | Missense | 7950 | CA119174 | | | | | | | | | | | | ✓ | | | | ✓ | | ✓ | | | | | | | | | PP1_Mod, PS3_Sup, PM2_Sup | 4 | VUS | P |
| 171605140G>T | 1440C>A | Asn480Lys | 3 | Missense | 7951 | CA119175 | | | | | ✓ | | | | | ✓ | ✓ | | | ✓ | | | | ✓ | | | | | | | | | PP1_Str, PS3_Mod, PS4_Mod, PP3, PM2_Sup | 10 | P | P |
| 171605140G>C | 1440C>G | Asn480Lys | 3 | Missense | | CA343723150 | ✓ | | | | | | | | | ✓ | | | ✓ | ✓ | | | | ✓ | | | | | | | | | PS1, PS3_Mod, PP1, PP3, PM2_Sup | 9 | LP | |
| 171605139G>A | 1441C>T | Pro481Ser | 3 | Missense | | CA1244022 | | | | | | | | | | | | | | | | | | ✓ | | | | | | | | | PM2_Sup | 1 | VUS | |
| 171605139G>T | 1441C>A | Pro481Thr | 3 | Missense | | CA16616748 | | | | | | | | | | | | | | ✓ | | | ✓ | ✓ | | | | | | | | | PP3, PS4_Sup, PM2_Sup | 3 | VUS | |
| 171605138G>A | 1442C>T | Pro481Leu | 3 | Missense | | CA343723135 | | | | | | | | | | ✓ | | | ✓ | ✓ | | | ✓ | ✓ | | | | | | | | | PS3_Mod, PP1, PP3, PS4_Sup, PM2_Sup | 6 | LP | |
| 171605138G>C | 1442C>G | Pro481Arg | 3 | Missense | | CA343723138 | | | | | | | | | | | | | | ✓ | | | | ✓ | | ✓ | | | | | | | PP3, PM2_Sup, PM5_Sup | 3 | VUS | |
| 171605097C>T | 1483G>A | Val495Ile | 3 | Missense | | CA343722912 | | | | | | | | | | | | | | | | | | ✓ | | | | | ✓ | | | | PM2_Sup, BS3_Mod | -1 | VUS | |
| 171605085T>A | 1495A>T | Ile499Phe | 3 | Missense | | CA343722844 | | | | | | | | | | ✓ | | ✓ | | | | | ✓ | ✓ | | | | | | | | | PS3_Mod, PP1_Mod, PS4_Sup, PM2_Sup | 6 | LP | |
| 171605084A>C | 1496T>G | Ile499Ser | 3 | Missense | | CA343722837 | | | | | | | | | | | | | | ✓ | | ✓ | | ✓ | | ✓ | | | | | | | PP3, PS3_Sup, PM2_Sup, PM5_Sup | 4 | VUS | |
| 171605076A>G | 1504T>C | Ser502Pro | 3 | Missense | | CA343722794 | | | | | | | | | | ✓ | | ✓ | | | | | | ✓ | | | | | | | | | PS3_Mod, PP1_Mod, PM2_Sup | 5 | VUS | |
